# Minimalistic Transcriptomic Signatures Permit Accurate Early Prediction of COVID-19 Mortality

**DOI:** 10.1101/2025.05.18.25327658

**Authors:** Rithwik Narendra, Emily C. Lydon, Hoang Van Phan, Natasha Spottiswoode, Lucile P. Neyton, Joann Diray-Arce, IMPACC Network, COMET Consortium, EARLI Consortium, Patrice M. Becker, Seunghee Kim-Schulze, Annmarie Hoch, Harry Pickering, Patrick van Zalm, Charles B. Cairns, Matthew C. Altman, Alison D. Augustine, Steve Bosinger, Walter Eckalbar, Leying Guan, Naresh Doni Jayavelu, Steven H. Kleinstein, Florian Krammer, Holden T. Maecker, Al Ozonoff, Bjoern Peters, Nadine Rouphael, Ruth R. Montgomery, Elaine Reed, Joanna Schaenman, Hanno Steen, Ofer Levy, Sidney A. Carillo, David Erle, Carolyn M. Hendrickson, Matthew F. Krummel, Michael A. Matthay, Prescott Woodruff, Elias K. Haddad, Carolyn S. Calfee, Charles R. Langelier

**Author notes:** Correspondence to: Charles R. Langelier, Department of Medicine, Division of Infectious Diseases University of California, San Francisco, 513 Parnassus Ave, Room HSE 401 San Francisco, CA 94122. These authors contributed equally (Rithwik Narendra, Emily C. Lydon).

## Abstract

**Background:** Predicting mortality risk in patients with COVID-19 remains challenging, and accurate prognostic assays represent a persistent unmet clinical need. We aimed to identify and validate parsimonious transcriptomic signatures that accurately predict fatal outcomes within 48 hours of hospitalization.

**Methods:** We studied 894 patients hospitalized for COVID-19 across 20 US hospitals and enrolled in the prospective Immunophenotyping Assessment in a COVID-19 Cohort (IMPACC) with peripheral blood mononuclear cells (PBMC) and nasal swabs collected within 48 hours of admission. Host gene expression was assessed by RNA sequencing, nasal SARS-CoV-2 viral load was measured by RT-qPCR, and mortality was assessed at 28 days. We first defined transcriptional signatures and biological features of fatal COVID-19, which we compared against mortality signatures from an independent cohort of patients with non-COVID-19 sepsis (n=122). Using least absolute shrinkage and selection operator (LASSO) regression in 70% of the COVID-19 cohort, we trained parsimonious prognostic classifiers incorporating host gene expression, age, and viral load. The performance of single and three-gene classifiers was then determined in the remaining 30% of the cohort and subsequently externally validated in an independent, contemporary COVID-19 cohort (n=137) with vaccinated patients.

**Results:** Fatal COVID-19 was characterized by 4189 differentially expressed genes in the peripheral blood, representing marked upregulation of neutrophil degranulation, erythrocyte gas exchange, and heme biosynthesis pathways, juxtaposed against downregulation of adaptive immune pathways. Only 7.6% of mortality-associated genes overlapped between COVID-19 and sepsis due to other causes. A COVID-specific three-gene peripheral blood classifier (*CD83, ATP1B2, DAAM2*) combined with age and SARS-CoV-2 viral load achieved an area under the receiver operating characteristic curve (AUC) of 0.88 (95% CI 0.82–0.94). A three-gene nasal classifier (*SLC5A5, CD200R1*, *FCER1A*), in comparison, yielded an AUC of 0.74 (95% CI 0.64-0.83). Notably the expression of *OLAH* alone, a gene recently implicated in severe viral infection pathogenesis, yielded an AUC of 0.86 (0.79–0.93). Both peripheral blood classifiers demonstrated comparable performance in vaccinated patients from an independent external validation cohort (AUCs 0.74– 0.80).

**Conclusions:** A three-gene peripheral blood signature, as well as *OLAH* alone, accurately predict COVID-19 mortality early in hospitalization, including in vaccinated patients. These parsimonious blood- and nasal-based classifiers merit further study as accessible prognostic tools to guide triage, resource allocation, and early therapeutic interventions in COVID-19.

## INTRODUCTION

The clinical course of SARS-CoV-2 infection is highly heterogeneous, ranging from minimal symptoms to fatal disease.^1,2^ Despite thousands of studies since the emergence of the virus in 2019^3^ and a growing understanding of the biological features underpinning severe COVID-19,^4–6^ clinicians still lack reliable prognostic assays to identify which patients will progress to critical illness or fatal disease. Accurate and timely severity prediction tools could improve clinical triage, optimize resource allocation, and have utility for predictive enrichment in clinical trials of novel therapeutics.^7–10^

Host factors including age^11^ and individual inflammatory responses are key determinants of disease severity and progression.^11–14^ Broadly available clinical laboratory tests, such as ferritin, D-dimer, lactate dehydrogenase, troponin, interleukin-6 and interleukin-8 have been used to risk stratify COVID-19 patients, but each biomarker individually has limited performance.^15,16^ Bioinformatic approaches attempted to integrate these laboratory values with clinical parameters, resulting in modest improvements in predictive ability.^17–20^ However, these studies have generally been single-institution studies, leveraging a small list of biomarkers, with concerns about model overfitting and lack of generalizability.^21^ No models have yet been implemented into an actionable, widely used prognostic tool in clinical practice.

In contrast to traditional protein-based biomarkers, host transcriptomic profiling offers a more comprehensive and less biased method for characterizing the host immune response to infection.^22^ Transcriptomic classifiers have increasingly shown promise in accurately diagnosing infection and predicting disease severity across a wide range of pathogens.^23–27^ A handful of early foundational studies, based on relatively small cohorts of ≤ 100 patients, have explored using host transcriptomic classifiers to predict COVID-19 severity.^28–31^

For instance, from a pre-existing panel of 29 genes, a six-gene prognostic classifier trained on blood transcriptomic data from non-SARS-CoV-2 viral infections was developed, which when tested in COVID-19 patients achieved AUCs ranging from 0.65-0.89.^28,29^ Similarly, another group repurposed a 10-gene sepsis mortality prediction score and found that it achieved an AUC of 0.86 in COVID-19 patients,^30^ and a third developed a 48-gene prognostic classifier that had an overall accuracy of 81%.^31^

While these early, important studies suggest that a transcriptomic COVID-19 severity classifier has potential, there remains an unmet need for a rigorously validated, clinically translatable mortality prediction tool, deployable at the time of hospitalization, with generalizability to diverse populations that include COVID-19-vaccinated individuals. Notably, all previously published classifiers rely on sizeable multi-gene combinations, while highly parsimonious (≤3 gene) classifiers have not yet been identified. Minimal gene expression models could enhance feasibility for clinical translation, reduce assay costs, and improve accessibility in resource-limited settings. Furthermore, compact gene signatures could be more readily incorporated into existing SARS-CoV-2 diagnostic platforms, facilitating rapid risk stratification at the time of diagnosis.

Here, we address this need by studying over 1000 COVID-19 patients enrolled in two cohorts across 20 hospitals in the United States. We identify single and three-gene signatures from peripheral blood and nasal swabs collected within 48 hours of hospital admission that accurately predict future COVID-19 mortality, including in vaccinated patients. We further demonstrate that incorporating patient age and SARS-CoV-2 viral load enhances prognostic ability of transcriptomic classifiers, offering a novel, translatable approach for early risk stratification in hospitalized COVID-19 patients.

## RESULTS

### Clinical and demographic features associated with fatal COVID-19

We analyzed 894 subjects enrolled in the Immunophenotyping Assessment in a COVID-19 cohort (IMPACC) who had peripheral blood and/or nasal swab samples collected at early timepoints in their hospitalization, as well as SARS-CoV-2 viral load measured in the upper airway (**Fig. 1**). We began by first evaluating the demographic and clinical features of fatal SARS-CoV-2 infection (**Table 1**). The overall mortality rate was 9.5%, and the survival group encompassed a wide range of illness severity based on maximal NIH respiratory ordinal score. Consistent with many prior studies, older age strongly correlated with mortality in COVID-19 patients from the IMPACC cohort (median 70.0 in mortality versus 58.0 in survival, P<0.001), as did higher viral load (median reverse transcriptase quantitative polymerase chain reaction (RT-qPCR) cycle threshold (CT) value of 25.5 in mortality vs 27.6 in survival, P=0.002). Most comorbidities that were evaluated were associated with mortality, including hypertension, diabetes, chronic lung disease, cardiovascular disease, chronic kidney disease, and malignancy. Therapeutically, steroid use was higher in patients who did not survive (81% vs 66%, P=0.005), though remdesivir use was similar (61% vs 63%, P=0.735). As subjects were enrolled between May 2020 and March 2021, this cohort was unvaccinated.

**Figure 1.**
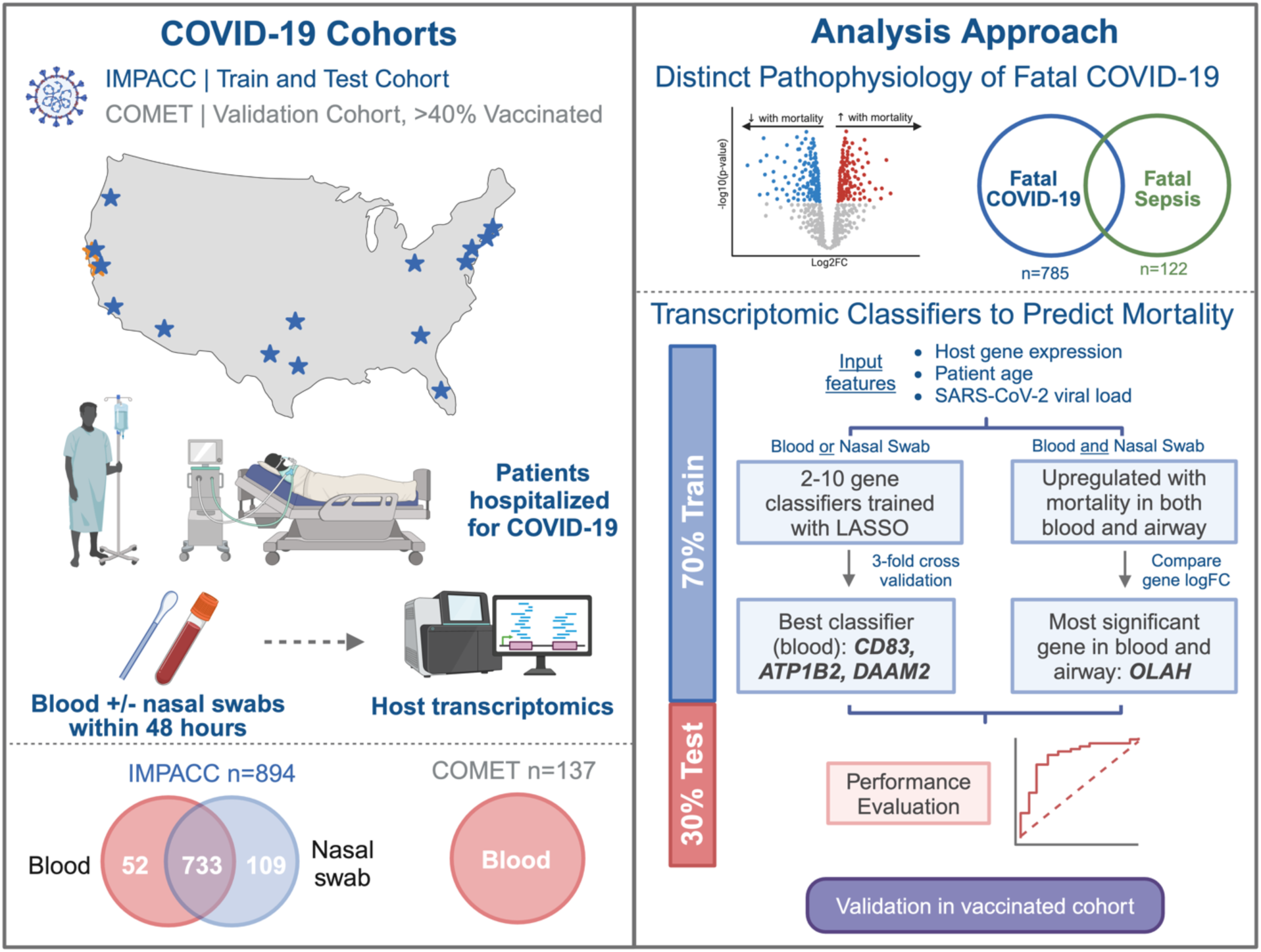
Overview schematic of study. This study evaluated 894 patients hospitalized with COVID-19 from the multi-center IMPACC cohort. Peripheral blood and nasal swab samples collected within 48 hours of hospitalization were utilized to evaluate host transcriptional signatures of mortality, which were then compared to sepsis mortality signatures. Parsimonious mortality prediction classifiers of varying lengths were then developed in train cohort (70%) and performance characteristics were assessed in the held-out test cohort (30%). Classifiers were then validated in an external cohort with vaccinated patients, and performance was compared to other larger classifiers published in the literature. IMPACC, ImmunoPhenotyping Assessment in a COVID-19 Cohort; COMET, COVID-19 Multi-Immunophenotyping Projects for Effective Therapies; QC, quality control; LASSO, least absolute shrinkage and selection operator; logFC = log gene expression fold change.

**Table 1:** Demographics of COVID-19 patients studied from the IMPACC cohort. Demographics are stratified by survival status. Mann-Whitney test was used for all continuous variables, and Fisher’s exact test was used for all categorical values. IQR, interquartile range; BMI, body mass index; ECMO, extracorporeal membrane oxygenation; HFNC, high-flow nasal cannula; CT, cycle threshold.

|  | Survival<br>(n=809) | Mortality<br>(n=85) | p-value |
| --- | --- | --- | --- |
| **Age, median (IQR)** | 58.0 (20.0) | 70.0 (16.0) | <0.001 |
| **Female sex, n (%)** | 311 (38%) | 24 (28%) | 0.064 |
| **Race, n (%)** |  |  | 0.124 |
| White | 387 (48%) | 55 (65%) |  |
| Black/African American | 188 (23%) | 12 (14%) |  |
| Asian | 33 (4%) | 4 (5%) |  |
| Other/Declined/Unknown | 201 (25%) | 14 (16%) |  |
| **Ethnicity, n (%)** |  |  | 0.013 |
| Non-Hispanic | 533 (66%) | 56 (66%) |  |
| Hispanic | 250 (31%) | 21 (25%) |  |
| Unknown | 26 (3%) | 8 (9%) |  |
| **Comorbidities, n (%)** |  |  |  |
| None | 56 (7%) | 3 (4%) | <0.001 |
| Hypertension | 448 (55%) | 62 (73%) | 0.002 |
| Diabetes | 286 (35%) | 41 (48%) | 0.019 |
| BMI > 30 | 449 (56%) | 43 (50.6%) | 0.949 |
| Chronic lung disease | 146 (18%) | 35 (41%) | <0.001 |
| Asthma | 126 (16%) | 9 (11%) | 0.222 |
| Chronic cardiac disease | 201 (25%) | 36 (42%) | <0.001 |
| Chronic kidney disease | 103 (13%) | 23 (27%) | <0.001 |
| Chronic liver disease | 42 (5%) | 4 (5%) | 0.847 |
| Organ Transplantation | 46 (6%) | 4 (5%) | 0.708 |
| HIV/AIDS | 9 (1%) | 1 (1%) | 0.957 |
| Malignancy | 68 (8%) | 13 (15%) | 0.035 |
| **Maximal O2 requirement n (%)** |  |  | <0.001 |
| Mechanical ventilated/ECMO | 65 (8%) | 29 (34%) |  |
| Non-invasive ventilation/HFNC | 128 (16%) | 29 (34%) |  |
| Supplemental O2 | 421 (52%) | 20 (24%) |  |
| None | 194 (24%) | 7 (8%) |  |
| Missing data | 1 (0%) | 0 (0%) |  |
| **Steroid use, n (%)** | 535 (66%) | 69 (81%) | 0.005 |
| **Remdesivir use, n (%)** | 510 (63%) | 52 (61%) | 0.735 |
| **Median SARS-CoV-2 PCR CT value (IQR)** | 27.6 (8.1) | 25.5 (6.7) | 0.002 |

### Early host transcriptional signatures of fatal COVID-19 exist in the blood and upper respiratory tract

We next evaluated the relationship between peripheral blood mononuclear cell (PBMC) gene expression profiles within 48 hours of hospital admission and COVID-19 mortality within 28 days (n=785). We identified 4189 differentially expressed genes (adjusted P value (P_adj_) <0.05), adjusting for sex and race (**Fig. 2A, Supp. Data 1A**). To explore their functions, we performed gene-set enrichment analysis (GSEA). Patients who died exhibited upregulation in genes related to erythrocyte gas exchange (e.g., *CA1* and *CA4*), heme biosynthesis (e.g., *HBA1*, *HBA2*, and *FECH*), neutrophil degranulation (e.g., *MPO* and *TNF*), among other pathways (**Fig. 2B, Supp. Data 1B)**. This was juxtaposed against downregulated expression of genes important for adaptive immunity, including B and T lymphocyte signaling (e.g*., CD22, CD79, CD96*, and *CD4*).

**Figure 2:**
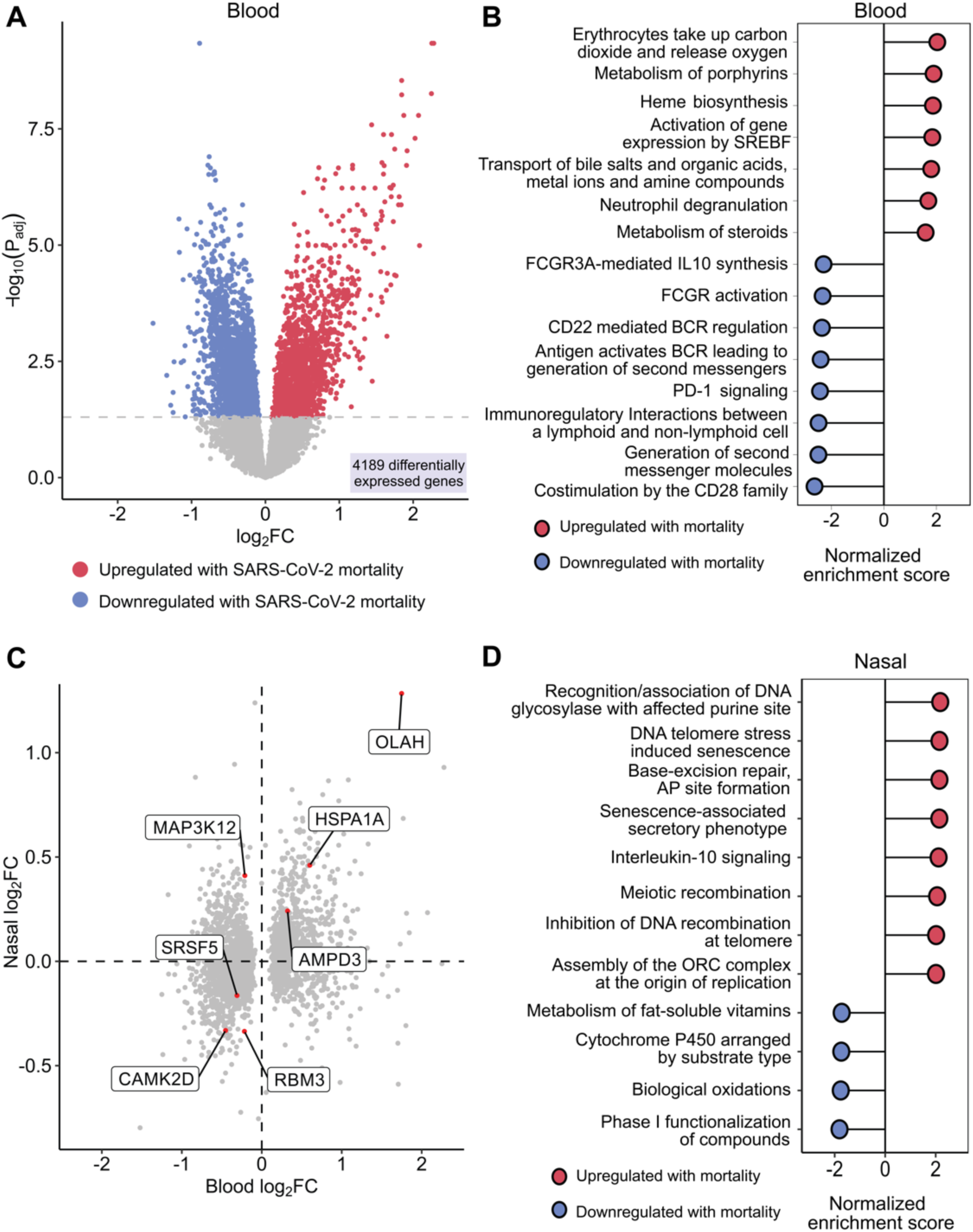
Early host gene expression signatures of mortality in the peripheral blood and upper airway. **A.** Volcano plot displaying the 4189 genes that were differentially expressed (DE) between survival and mortality in the peripheral blood of COVID-19 patients, using a Benjamini-Hochberg adjusted p value of 0.05. **B.** Gene set enrichment analysis (GSEA) demonstrating statistically significant pathways associated with mortality based on Benjamini-Hochberg adjusted p-value in the peripheral blood (red = upregulated with mortality, blue = downregulated with mortality). **C.** Log-log plot demonstrating the 7 genes that were DE in both peripheral blood and nasal swab samples. log_2_FC = base 2 logarithm of fold change. **D.** GSEA demonstrating the differentially expressed mortality pathways in nasal samples (red = upregulated pathways, blue = downregulated pathways).

We performed a similar analysis of transcriptomic data derived from nasal swabs collected within 48 hours of hospital admission (n=842). A host signature of mortality was also present in the upper respiratory tract, although differential gene expression was more subtle, with only 53 genes significantly associated with mortality (**Supp. Fig. 1A, Supp. Data 2A)**. Of these, seven were consistently differentially expressed across both the peripheral blood and the upper airway, with *OLAH*, which encodes oleoyl-ACP hydrolase, most strongly upregulated with mortality in both nasal swab and PBMC samples (**Fig. 2C**). In the upper airway, mortality was associated with nucleic acid repair, cellular senescence and IL-10 signaling pathways (**Fig 2D, Supp. Data 2B**).

### The transcriptional signature of fatal COVID-19 has unique features compared to fatal sepsis

Understanding whether the host response leading to death in COVID-19 is distinct from or shared from other forms of sepsis could provide insights into disease-specific mechanisms or risk stratification strategies. We therefore sought to determine whether host transcriptional signatures of mortality early in hospital admission were similar or different between patients hospitalized for COVID-19 versus sepsis due to other causes. To address this question, we analyzed peripheral blood RNA-seq data from 122 patients hospitalized for microbiologically confirmed sepsis prior to the COVID-19 pandemic, a cohort which had a 34.4% mortality rate and a predominance of bacterial infections (**Figure 3A, Table S1**).^32,33^ We identified a distinct host signature of sepsis mortality characterized by 1246 differentially expressed genes (**Supp. Data 3A)**.

**Figure 3:**
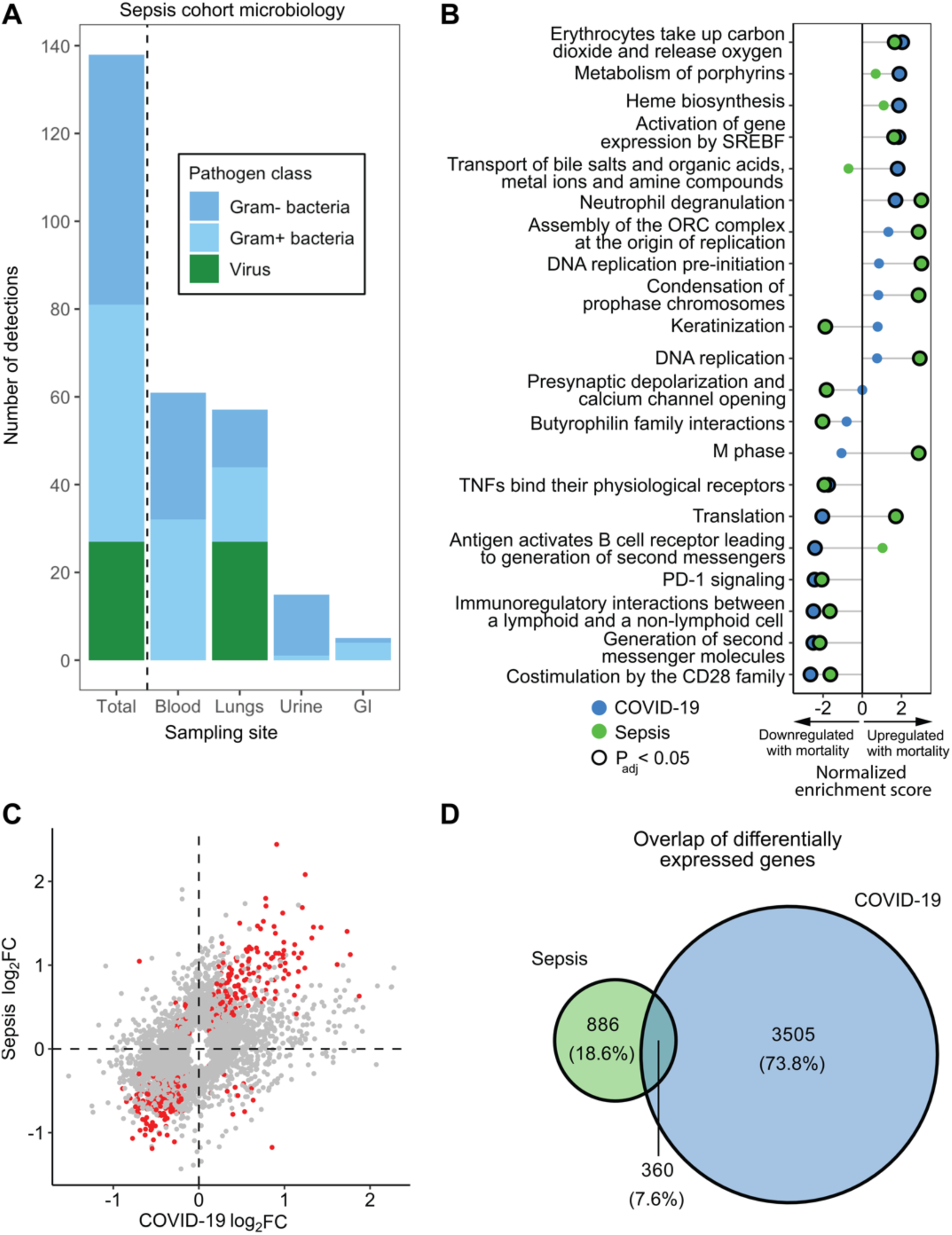
Comparison of mortality signatures between COVID and non-COVID sepsis. **A.** Microbiology of the non-COVID sepsis cohort (n=122), stratified by sampling site and pathogen category. The total bar on the left shows the summation of the right sided bars, and the number of detections exceeds the number of patients as some patients had multiple pathogen classes detected. **B.** Gene set enrichment analysis (GSEA) of COVID-19 mortality (blue circles) overlaid that of sepsis mortality (green circles). Circles outlined in black are statistically significant based on Benjamini-Hochberg adjusted P values (P_adj_). **C.** Log-log plot of differentially expressed genes in COVID-19 and sepsis mortality, with genes statistically significant in both (based on P_adj_) highlighted in red. **D.** Venn-diagram highlighting the limited overlap (7.6%) between DE genes in COVID-19 and sepsis.

At the biological pathway level, GSEA demonstrated that fatal COVID-19 and non-COVID sepsis were both characterized by increased expression of neutrophil degranulation genes and downregulation of T cell signaling genes (**Fig. 3B, Supp. Data 3B**). Fatal COVID-19, however, was uniquely characterized by impaired expression of genes related to B cell signaling and translation, and increased expression of genes functioning in heme biosynthesis. Differences between fatal COVID-19 and non-COVID sepsis were even more apparent at the individual gene level (**Fig. 3C**), with only 360 (7.6%) of mortality-associated genes shared between groups (**Fig. 3D**). Taken together, these results demonstrated that fatal SARS-CoV-2 infection has unique transcriptional changes compared to sepsis caused by other pathogens, suggesting that accurate prognostic assessment for COVID-19 warrants a classifier specifically trained on these distinctive COVID-19 mortality signatures rather than relying on classifiers developed for other critical illnesses.

### Parsimonious host-viral classifiers accurately predict COVID-19 mortality

Given the striking transcriptomic signature of COVID-19 mortality, we next sought to build prognostic classifiers based on gene expression measured within the first 48 hours of hospitalization. To maximize potential for future clinical translation, we sought to identify parsimonious feature sets of ≤ 10 genes. For derivation of the classifiers, we divided the cohort into training (70% of patients) and test sets (30%) (**Fig. 1**). Given that age and SARS-CoV-2 viral load (RT-qPCR CT value) are well-established risk factors for fatal COVID-19 and readily obtainable from all hospitalized patients, we included both as additional parameters in the models.

We first used least absolute shrinkage and selection operator (LASSO) regression to build 2-10 gene peripheral blood classifiers within the training set **(Table S2)**. These gene sets were combined in a logistic regression model with age and SARS-CoV-2 CT value, and performance distribution was assessed using a three-fold repeated random partitioning approach **(Fig. 4A)**.

**Figure 4.**
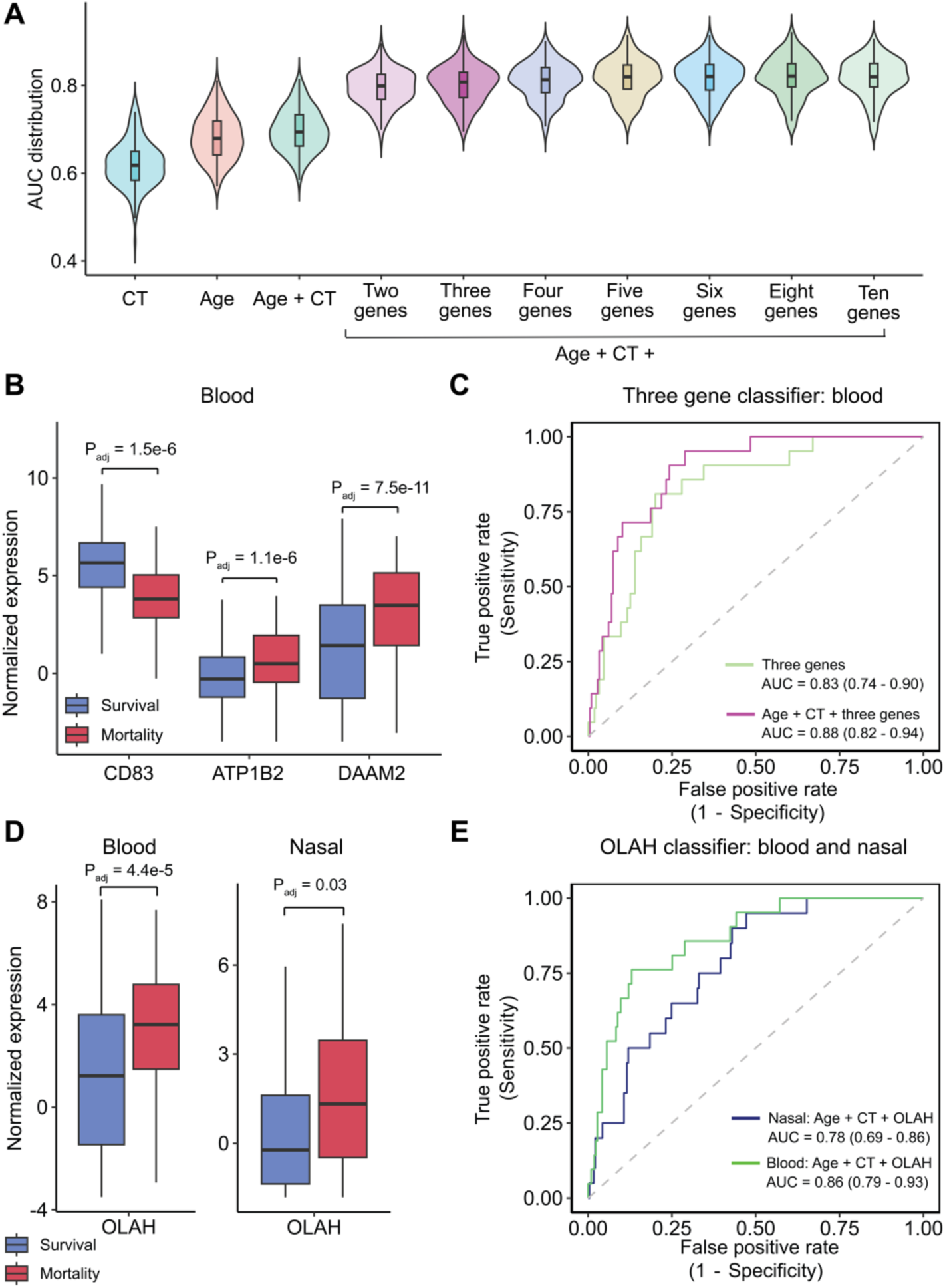
Parsimonious host-viral classifiers predict COVID-19 mortality. **A.** Violin plots showing the area under the curve (AUC) distribution for each of the peripheral blood candidate classifiers, evaluated in the training cohort. Violin plots showing the performance of SARS-CoV-2 cycle threshold (CT), age, and age + CT are displayed to the left. **B.** Boxplots comparing the log_2_ counts per million normalized gene expression of the three genes in the optimally performing classifier between survival (blue) and mortality (red) in the full cohort. **C.** Receiver operating characteristic (ROC) curves for the three-gene classifier, alone and with the addition of age + CT, as evaluated in the test set (AUC +/- 95% confidence interval). **D.** Boxplots comparing the log_2_ counts per million normalized gene expression for *OLAH* in blood (left) and nasal swab (right) between survival (blue) and mortality (red) for the full cohort. **E.** ROC curves for OLAH classifiers with the addition of age and CT value in the test set (AUC +/- 95% confidence interval). All P-values (P_adj_) shown were adjusted for multiple comparisons using the Benjamini-Hochberg method.

We found that classifier performance plateaued at a classifier size of three genes, with the combination of *CD83, ATP1B2*, and *DAAM2* performing as well as the larger gene sets (**Fig. 4B**). *CD83* plays a role in the activation of B cells and dendritic cells;^34,35^ *ATP1B2*, a component of sodium-potassium pumps that are important for maintaining endothelial integrity;^36^ *DAAM2* regulates the Wnt signaling pathway, thereby influencing cell fate.^37^ When tested in the held-out 30% validation set, this three-gene classifier achieved an AUC of 0.88 (95% CI 0.82-0.94) (**Fig. 4C)**.

Using the same methodology, we derived classifiers using nasal swab transcriptomic data in the training set (**Fig. S1B, Table S3**). However, the best performing three-gene set (*SLC5A5, CD200R1*, *FCER1A,* **Fig. S1C**) only achieved an AUC of 0.74 (95% CI 0.64-0.83) when evaluated on the held-out test set (**Fig. S1D)**. Given that *OLAH* expression was conspicuously amplified in fatal COVID-19 both in the upper respiratory tract and blood (**Fig. 3C**, **Fig 4D**), and because *OLAH* was recently implicated in the pathogenesis of severe viral pneumonia,^38^ we also evaluated its performance in a single-gene classifier. When assayed in the blood, in combination with age and SARS-CoV-2 CT value, *OLAH* remarkably achieved an AUC of 0.86 (0.79-0.93) **(Fig 4E)**. When assessed in the upper airway, an *OLAH* prognostic classifier achieved an AUC of 0.78 (0.69-0.86) **(Fig 4E)**. Taken together, these findings demonstrated that 1-3 gene parsimonious classifiers from either blood or nasal swab samples can accurately predict future COVID-19 mortality.

### Validation in an independent cohort with vaccinated COVID-19 patients

We next explored the extent to which our findings were generalizable. To that end, we leveraged the COVID-19 Multi-Immunophenotyping Projects for Effective Therapies (COMET) cohort, which enrolled COVID-19 positive patients (PBMC, n=137) at two hospitals through 2023 and notably included 55 (40.1%) vaccinated patients (**Table S4**).^39^ Differential expression analysis yielded 769 DE genes, confirming a robust peripheral blood signature of mortality in this validation cohort **(Fig. 5A, Supp. Data 4A**). GSEA demonstrated that neutrophil degranulation and erythrocyte transport of oxygen and carbon dioxide remained two of the most significantly upregulated pathways with mortality, but showed a notable absence of significantly downregulated adaptive immunity pathways (**Fig. S2, Supp. Data 4B)**.

**Figure 5.**
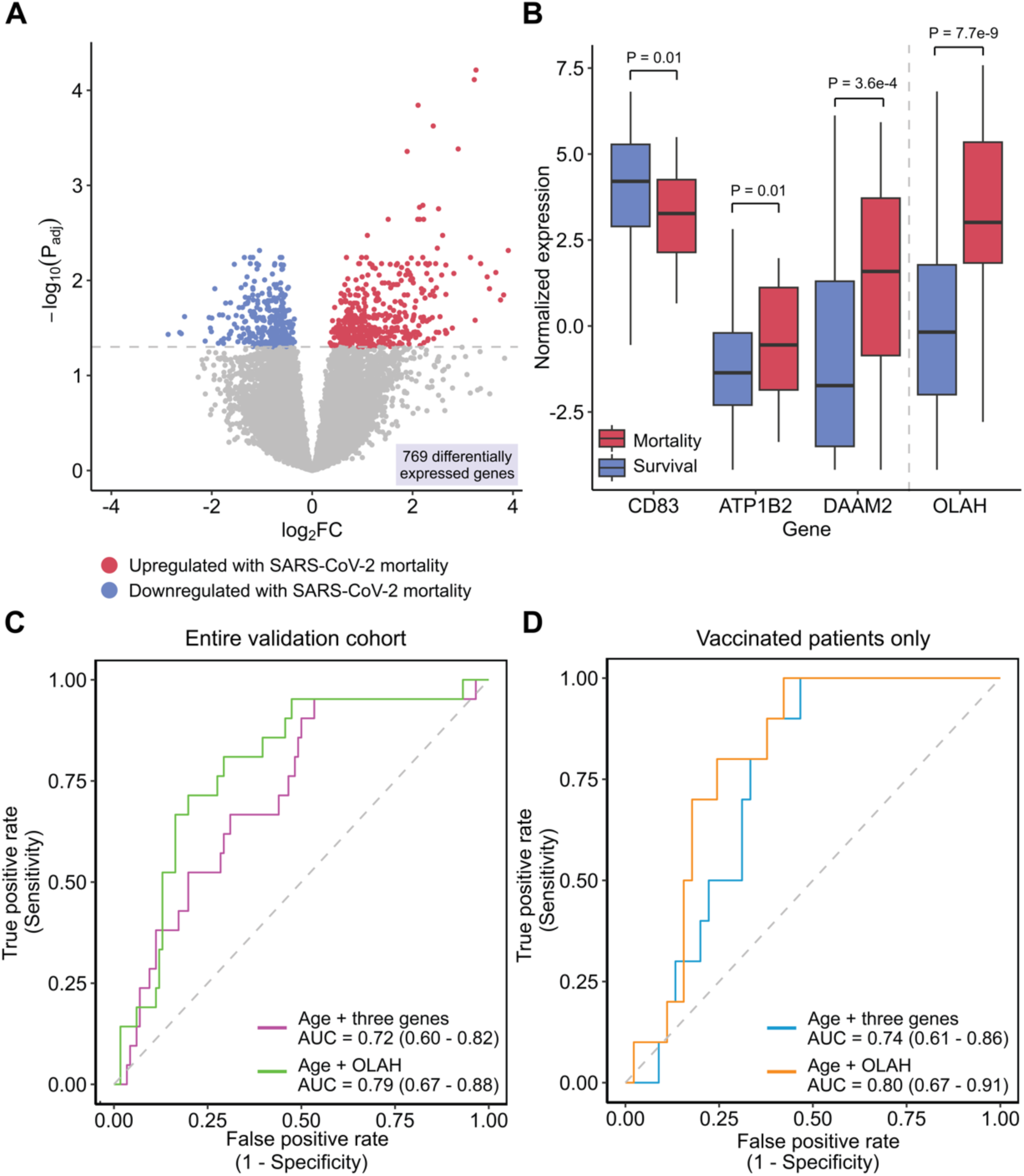
Validation of COVID-19 mortality signature and host-viral classifiers in an independent cohort including vaccinated patients. **A** Volcano plot demonstrating the 769 differentially expressed genes between mortality and survival in the validation cohort (red = upregulated with mortality, blue = downregulated with mortality), using a Benjamini-Hochberg adjusted p-value of 0.05. **B.** Boxplots comparing the log_2_ counts per million normalized gene expression of the three-gene classifier genes (*CD83*, *ATP1B2*, *DAAM2*) and single-gene OLAH classifier between survival (blue) and mortality (red) in the external validation data set. **C.** Performance of three-gene classifier (purple) and single-gene classifier (green) with the added feature of age in the validation cohort. Area under the curve (AUC) listed as value +/- 95% confidence interval**. D**. Performance of three-gene classifier (orange) and single-gene classifier (teal) with the added feature of age in the validation cohort in vaccinated patients only.

Despite some minor differences at the biological pathway level, the expression of *OLAH* as well as *CD83, ATP1B2*, and *DAAM2* differed significantly (P<0.05) based on mortality in the validation cohort (**Fig. 5B**). Because SARS-CoV-2 CT value was not available on COMET patients, we tested the genes in combination with just age. Using five-fold cross validation followed by out-of-fold AUC calculation, *OLAH* achieved an AUC of 0.79 (0.67 - 0.88), and the three-gene classifier an AUC of 0.72 (0.60 - 0.82) (**Fig. 5C**). When repeating this for vaccinated patients, the three-gene and *OLAH* classifiers performed equally well, if not better, at predicting mortality in the vaccinated subset (**Fig. 5D**). Collectively, these findings demonstrated that these transcriptomic classifiers remained capable of predicting mortality in an independent cohort inclusive of vaccinated individuals.

### Parsimonious single and three-gene prognostic classifiers perform comparably to a larger multi-gene classifier

Finally, we sought to compare the results of our single-gene *OLAH* classifier and our three-gene classifier to a previously published six-gene classifier (*HK3, LY86, TGFB1, DEFA4, BATF,* and *HLA-DPB1)* that was developed in non-COVID-19 viral infections and previously tested in COVID-19 patients.^28^ These six genes trained and tested in the IMPACC cohort yielded an AUC of 0.75, which improved to 0.88 after including age and CT value (**Fig. S3)**. Notably, the AUCs and confidence intervals of the published six-gene was comparable to our integrated three-gene and single-gene *OLAH* classifier, indicating that one and three-gene classifiers can perform comparably to larger size classifiers, and that adding age and viral load can boost the performance of existing classifiers for COVID-19 risk stratification.

## DISCUSSION

In a large, prospective, multi-center cohort, we find that either a single gene, *OLAH,* or a combination of three genes, *CD83, ATP1B2*, and *DAAM2,* accurately predicts 28-day mortality in hospitalized COVID-19 patients, including those who have been vaccinated. We build on extensive foundational studies establishing clinical and biological risk factors for severe COVID-19^12,40–42^ by characterizing early host transcriptional determinants of survival versus death in comparison to hospitalized patients with non-COVID-19 sepsis. We then leverage these findings to build parsimonious host-based classifiers from both blood and nasal swab samples that can be readily adapted to existing nucleic acid amplification platforms for clinical deployment.

The three-gene classifier achieved an AUC of 0.88 (0.82-0.94) and remarkably, the single-gene *OLAH* classifier achieved an AUC of 0.86 (0.79-0.93) in the blood and 0.78 (0.69-0.86) in nasal swab samples. Similar performance was observed in an independent validation cohort, which included vaccinated patients, and mortality prediction was similar when stratifying by vaccinated status. Several prior studies, representing important early contributions, developed severity or mortality prediction classifiers for COVID-19^28–31,43^. Each, however, was limited by small sample sizes, single-institution cohorts, development in non-COVID-19 populations, and testing in unvaccinated patients. Additionally, these classifiers incorporated anywhere from six to 48 genes, whereas we identified single and three-gene classifiers that achieved equivalent performance in head-to-head comparisons.

While severe COVID-19 is less common now than in the beginning of the pandemic, there is still considerable mortality with each wave.^44^ Simple, rapid prognostic tests for COVID-19 could not only aid in clinical triage and resource allocation during surges but could also identify high-risk patients who may benefit from early targeted interventions.^8,24^ Reducing the number of targets substantially decreases the technical and computational complexity of host gene expression tests, as well as their cost, making clinical implementation more feasible, especially in resource limited settings where risk stratification may be disproportionately needed.^45^ The strong performance of *OLAH* in both blood and nasal swabs samples makes it particularly attractive for translation, as this gene could be readily incorporated into existing nasal swab SARS-CoV-2 diagnostic assays to additionally enable prognostication.

Beyond their prognostic potential, each of the classifier genes we report may contribute to the pathophysiology of severe COVID-19, as each has been previously linked to COVID-19 in the literature.^46–48^ *CD83*, a well-established regulator of immune responses that promotes T and B cell maturation, was downregulated in fatal COVID-19,^49^ consistent with impaired adaptive immunity. *ATP1B2* encodes a regulatory subunit of the sodium/potassium-transporting ATPase pump, and its dysregulation may disrupt vascular endothelial^36,50^ and alveolar epithelial integrity^51^, promoting capillary leak and acute respiratory distress syndrome. *DAAM2* negatively regulates Wnt signaling,^52^ a pathway implicated in the activation of inflammatory macrophages,^53^ angiogenesis,^54^ endothelial integrity,^55^ and fibrosis^56^. Elevated *DAAM2* has been linked with vascular disorders of pregnancy,^37,57^ suggesting it may also contribute to the widespread vascular dysfunction observed in fatal COVID-19.^58^

OLAH (oleoyl-acyl-carrier-protein hydrolase), an enzyme involved in fatty acid biosynthesis, was recently implicated in the pathogenesis of life-threatening respiratory viral infections.^38^ *OLAH*-knockout mice demonstrate protection against severe influenza infection, reduced inflammatory damage, and improved control of viral replication, outcomes attributed to modulating lipid mediators of inflammation.^38^ Importantly, *OLAH* expression was found to be increased across patients with severe influenza virus, RSV and SARS-CoV-2 infection,^38^ suggesting that an *OLAH* prognostic classifier may be generalizable across a diversity of respiratory viral infections. Here, we independently validate the association of both airway and peripheral blood *OLAH* expression with COVID-19 severity, and provide the first assessment of its performance as a prognostic biomarker in two large cohorts of hospitalized patients.

A growing body of literature demonstrates that severe COVID-19 is driven by a profoundly dysregulated host immune response to the virus, characterized by excessive innate inflammation and impaired adaptive immunity.^5,59^ Hyperactive neutrophils and macrophages contribute to cytokine release, complement activation, endothelial damage, and vascular thrombosis, while impaired lymphocyte responses delay viral clearance and increase vulnerability to secondary infections.^60–66^ In our study, the systemic host signature of fatal COVID-19 mirrored many of the same pathways described previously, including increased neutrophil degranulation, decreased production of the anti-inflammatory cytokine IL-10, and downregulated T-cell and B-cell signaling – highlighting that this aberrant immune signaling begins early in the course of illness. In addition, we noted increased systemic expression of erythrocyte gas exchange and heme biosynthesis genes in fatal cases, likely a compensatory mechanism for severe hypoxemia.^67,68^

The host signature of mortality in the upper respiratory tract, in contrast, was dominated by upregulation of DNA repair and senescence pathways, potentially reflecting heightened cellular stress and direct damage at the site of viral entry.^69,70^ Intriguingly, IL-10 signaling was upregulated in the upper respiratory tract but downregulated systemically, suggesting a localized attempt to control inflammation that fails to extend to the systemic immune response.^71,72^

Many of the pathways enriched in fatal COVID-19 have also been described in fatal sepsis.^73,74^ Prior studies have suggested that critically ill patients may follow a common mortality trajectory, with one study reporting that COVID-19 and non-COVID-19 patients admitted to the ICU for greater than seven days were almost transcriptionally indistinguishable.^75^ While we did find overlapping mortality-associated signaling pathways when comparing our GSEA results against those from a cohort of primarily bacterial sepsis patients (e.g., upregulated neutrophil degranulation, downregulated T cell signaling), more than 90% of mortality-associated genes differed between COVID-19 and sepsis. Fatal COVID-19 was uniquely characterized by decreased expression of B-cell signaling genes and enrichment of heme biosynthesis pathways. These findings suggest that while mortality pathways may eventually converge, pathogen-specific mortality signatures are prominent early in the course of severe disease – an important consideration for developing accurate early mortality prediction tools.

Strengths of our study include a large sample size, a multi-center design, incorporation of multiple different sample types, and a rigorous informatics approach for classifier development. However, our study also has limitations. The primary cohort was recruited before COVID-19 vaccines became widely available; however, we found that our classifiers performed equally well in a cohort inclusive of vaccinated patients. Additionally, our cohorts consisted solely of symptomatic hospitalized patients, leaving uncertainty about whether the classifiers would maintain their performance in outpatient settings.

Taken together, we present a comprehensive transcriptomic characterization of fatal COVID-19, illuminating key pathways driving severe outcomes and developing parsimonious blood- and nasal swab-based classifiers accurately predict mortality COVID-19 early in illness. Moving forward, next steps involve further validation of classifiers across a broader spectrum of disease severity, translation onto a point-of-care clinical platform, and real-world assessment of their impact on patient management and outcomes.

## METHODS

### Sex as a biological variable

Sex as a biological variable. Our study examined male and female participants. Classifiers were designed to predict 28-day mortality, an outcome that did not differ by sex in our study.

### Study cohorts and design

This study primarily leveraged the Immunophenotyping Assessment in a COVID-19 Cohort (IMPACC) observational cohort, which enrolled a total of 1164 patients hospitalized for COVID-19 from 20 different US hospitals^76,77^. Biological sample collection, processing, and multi-modal immune profiling followed a standard protocol utilized at core laboratories and by every participating academic institution^76,77^. The Department of Health and Human Services Office for Human Research Protections determined that the IMPACC study protocol met criteria for a public health surveillance exception [45CFR46.102(l)(2)], and the study was approved by each institutional review board (IRB) through this exception (12 sites) or by pre-approved biobanking protocols (3 sites).

For our primary analyses, we included all IMPACC participants who met the following inclusion criteria: 1) had at least one nasal swab or peripheral blood mononuclear cells (PBMC) collected within the first 48 hours of hospital admission for RNA-seq, and 2) had an admission SARS-CoV-2 viral load measured by either RT-qPCR or RNA-seq. Samples that failed RNA-seq quality control standards (described below) were removed, ultimately leaving 894 total patients included. For subjects with multiple available samples that met these criteria, only the earliest nasal swab and PBMC sample were retained. Both PBMC transcriptomic data and SARS-CoV-2 viral load measurements were available for 785 IMPACC participants. Nasal transcriptomic data and SARS-CoV-2 viral load measurements were available for 842 IMPACC participants.

Two external cohorts were also studied. We validated our classifiers in the COVID-19 Multi-immunophenotyping projects for Effective Therapies (COMET) cohort of patients hospitalized for COVID-19 at UCSF and Zuckerberg San Francisco General (ZSFG) hospitals between 2020 and 2023 (n=137, UCSF IRB Protocol #20-30497).^39^ In addition, we compared our findings against critically ill adults with sepsis due to causes other than COVID-19 who were enrolled in the Early Assessment of Renal and Lung Injury (EARLI) cohort between 2013 and 2018 at UCSF and Zuckerberg San Francisco General (ZSFG) hospitals in San Francisco, CA, USA (n=122, UCSF IRB Protocol #10-02852).^78^

### Standardization of SARS-CoV-2 viral load measurements

Viral load was measured from nasal swab samples either using either SARS-CoV-2 RT-qPCR (CT) or RNA-seq (reads-per-million, rPM), which were highly correlated (P < 2e-16, **Fig. S4**). Because some subjects did not RT-qPCR performed, we imputed CT values from rPM using a regression model generated on subjects who had both samples available. Specifically, we fit a robust regression model using the lmrob function from the R package robustbase^79^ on the log-transformed rpM values and CT data using the formula CT ∼ ln(rpM+1) (**Fig. S4**). If subjects had both viral load measurement types available, the CT measurement was used.

### Host gene expression analysis

RNA-seq and alignment against the host transcriptome was performed as previously described,^76,77^ and the deidentified, quality-controlled raw gene count files and metadata were obtained from the IMPACC study. We retained samples with at least 10,000 genes and retained protein-coding genes that had a minimum of 10 counts in at least 20% of the samples. Differential expression analyses were performed comparing mortality and survival using the R package limma using quantile normalization and the voom method,^80,81^ and age and sex were included as covariates. The eBayes function with default parameters was employed to compute empirical Bayes statistics and calculate the P values, correcting for multiple testing with Benjamini-Hochberg method. Adjusted P values <0.05 were considered significant.

Gene set enrichment analysis (GSEA) was performed using the R package fgsea,^82^ applying REACTOME pathways with a minimum size of 5 genes and a maximum size of 500 genes.^83^ All genes from the limma differential expression analyses were included as input, pre-ranked in descending order using the equation -log_10_(adjusted P value) * sign(log_2_(fold change)). Pathways with adjusted p values <0.05 were considered significant.

Identical differential expression and pathway analyses were performed on PBMC and nasal swab RNA-seq data from IMPACC.

### Comparison of COVID-19 and sepsis mortality

We compared the biological pathways enriched in COVID-19 mortality to those enriched in sepsis mortality, leveraging whole blood RNA-seq data from patients with sepsis enrolled in the EARLI cohort.^78^ Differential expression and pathway analyses comparing survival and mortality in EARLI were performed in the exact same manner as with IMPACC. The top six up- and down-regulated pathways for COVID-19 and sepsis were selected for visualization. If a pathway did not have GSEA results, the enrichment score was set to zero.

### Development of parsimonious mortality prediction classifiers

PBMC and nasal swab mortality prediction classifiers were generated separately. For each, subjects were first randomly divided into a train set (70%) and test set (30%). Input features for the classifier models included gene expression data, age, and SARS-CoV-2 viral load data. For the train set, we filtered for protein-coding genes with log_2_(fold change) >1 or <-1 and adjusted P value <0.05 based on the differential expression analysis described previously, in addition to employing standard filtering for protein-coding genes with at least 10 counts in at least 20% of the samples. This gene filter was subsequently applied to the test set. We performed variance-stabilizing transformation on the train set using the R package DESeq2.^84^ These dispersion estimates were then applied to the test set. We standardized gene expression, age, and SARS-CoV-2 viral load features in the train set using the caret R package^85^ and applied these standardization parameters to the test set.

The train set was used to identify the optimal classifier, and the test set was used to evaluate performance of this classifier. First, the train set was divided into five folds for the feature selection step, maintaining a relatively even distribution of mortality across each fold. We methodically iterated through multiple candidate gene sets of varying classifier lengths (ranging from n=2 genes to n=10 genes for PBMC and n=2 genes to n=4 genes for nasal swab, as the latter had far fewer genes that passed filtering). Specifically, for each n, we employed the Least Absolute Shrinkage and Selection Operator (LASSO) model in the glmnet R package,^86,87^ performing five-fold cross validation by training the model on four of the five folds and testing on the remaining fold, ultimately yielding five gene lists for each n length. Age and SARS-CoV-2 viral load which were included as additional features in each of the classifiers.

We next tested the performance of each candidate classifier (five classifiers for each gene length n) by using repeated random partitioning. Specifically, we performed 50 iterations of randomly splitting the data into three folds using the createFolds() function from the caret package^85^ and used logistic regression with the classifier features to obtain a distribution of 150 area under the curve (AUC) values for each feature set using the pROC R package.^88^ The AUC distribution was plotted for each of the best performing classifiers of length n (i.e., those with the highest average AUC), as well as for classifiers only incorporating viral load, age, and viral load + age. From these, the classifier with the fewest genes where the average AUC plateaued was chosen as the final classifier.

Finally, we evaluated the performance of these classifiers and a classifier consisting of *OLAH* expression, viral load, and age, on the held out test set. We fit a logistic regression model on the full train set and made predictions on the test set. We generated a ROC curve and calculated AUC and confidence intervals using the pROC package, as described above. We computed a 95% confidence interval for our AUC value using the ci.auc (method = “bootstrap”, boot.n = 5000, boot.stratified = TRUE) function.

### External validation of mortality classifiers

We externally validated our peripheral blood COVID-19 mortality prediction classifiers in the COMET cohort, which included vaccinated patients. We included all COMET subjects that were not co-enrolled in IMPACC and had PBMC available on the day of study enrollment (n=137). Gene expression quality control, filtering, normalization, and transformation were done with identical methods as in IMPACC. Differential expression and GSEA were conducted similarly, controlling for an additional covariate of batch. We evaluated two classifiers with different model features: three classifier genes and age, and *OLAH* and age. SARS-CoV-2 viral load information was not available in COMET. We re-trained five logistic regression models using these input features in COMET by employing the same five-fold cross validation as described previously to generate ROC curves and AUC metrics. We then computed the AUC values on the out-of-fold predictions and bootstrapped as described previously.

### Comparison to an existing mortality classifier

We compared our peripheral blood *OLAH* classifier and three-gene classifier to a previously reported six-gene mortality classifier originally developed in patients with sepsis^28^. In the IMPACC train set, we trained a logistic regression model using the six genes (*HK3, LY86, TGFBI, DEFA4, BATF, and HLA-DPB1*), with and without the addition of age and SARS-CoV-2 viral load as input parameters. We evaluated our models in the test set and generated ROC curves, using the same methodology described above for three-gene signature and *OLAH* classifiers.

## Data and code availability

Data used in this study is available at ImmPort Shared Data under the accession number <u>SDY1760</u> and in the NLM’s Database of Genotypes and Phenotypes (dbGaP) under the accession number <u>phs002686.v2.p2</u>. All code is deposited in the following Bitbucket repository: https://bitbucket.org/kleinstein/impacc-public-code/src/master/mortality_prediction_manuscript/.

## Data Availability

Data used in this study is available at ImmPort Shared Data under the accession number SDY1760 and in dbGAP under the accession number phs002686.v2.p2.

https://bitbucket.org/kleinstein/impacc-public-code/src/master/mortality_prediction_manuscript/

## Author contributions

E.C.L., R.N., H.V.P. and C.R.L. conceived the idea for the project. M.C.A., S.E.B., W.E., N.D.J., and S.K-S. generated the data. The I.N. and C.C. contributed to cohort design, participant enrollment, sample collection, data generation and/or data quality assurance. R.N., E.C.L., H.V.P., N.S., L.P.N., and A.H. analyzed the data. R.N., E.C.L., H.V.P., N.S., L.P.N., J.D-A., P.M.B., S.K-S., A.H., H.P. P. vZ., C.B.C., M.C.A. A.D.A., S.E.B., W.E., L.G. N.D.J., S.H.K., F.K., H.T.M., A.O. B.P., N.R., R.R.M., E.R., J.S., H.S., O.L., S.A.C., D.E., C.H., M.K., M.A.M., P.W., E.K.H., C.S.C., and C.R.L. provided input on analyses and findings. R.N., E.C.L., H.V.P. and C.R.L. wrote the manuscript. R.N., E.C.L., H.V.P., N.S., L.P.N., J.D-A., P.M.B., S.K-S., A.H., H.P. P. vZ., C.B.C., M.C.A. A.D.A., S.E.B., W.E., L.G. N.D.J., S.H.K., F.K., H.T.M., A.O. B.P., N.R., R.R.M., E.R., J.S., H.S., O.L., S.A.C., D.E., C.H., M.K., M.A.M., P.W., E.K.H., C.S.C., and C.R.L. reviewed and edited the manuscript.

## Acknowledgements

We thank the participants of the study for their voluntary enrollment and contribution of samples for this work. See the supplement for details on the IMPACC Network. We acknowledge the assistance of the following individuals: Sanya Thomas, Mitchell Cooney, Shun Rao, Sofia Vignolo, and Elena Morrocchi (all from the CDCC); Arash Naeim, Marianne Bernardo, Sarahmay Sanchez, Shannon Intluxay, Clara Magyar, Jenny Brook, Estefania Ramires-Sanchez, Megan Llamas, Claudia Perdomo, Clara E. Magyar, and Jennifer A. Fulcher (all from the David Geffen School of Medicine at UCLA); members of the UCLA Center for Pathology Research Services and the Pathology Research Portal; M. Catherine Muenker, Dimitri Duvilaire, Maxine Kuang, William Ruff, Khadir Raddassi, Denise Shepherd, Haowei Wang, Omkar Chaudhary, Syim Salahuddin, John Fournier, Michael Rainone, and Maxine Kuang (all from the Yale School of Medicine). We thank the leadership of Boston Children’s Hospital including Drs. Wendy Chung, Gary Fleisher and Kevin Churchwell for their support for the Precision Vaccines Program. Dr. Augustine’s and Becker’s co-authorship of this report does not necessarily represent the official views of the National Institute of Allergy and Infectious Diseases, the National Institutes of Health or any other agency of the United States Government. This work was supported by the United States National Institutes of Health: (5R01AI135803-03, R35HL140026, 5U19AI118608-04, 5U19AI128910-04, 4U19AI090023-11, 4U19AI118610-06, R01AI145835-01A1S1, 5U19AI062629-17, 5U19AI057229-17, 5U19AI125357-05, 5U19AI128913-03, 3U19AI077439-13, 5U54AI142766-03, 5R01AI104870-07, 3U19AI089992-09, 3U19AI128913-03, 5T32DA018926-18, and K0826161611); NIAID, NIH (3U19AI1289130, U19AI128913-04S1, and R01AI122220); NCATS (UM1TR004528), the National Science Foundation (DMS2310836) and the Chan Zuckerberg Biohub San Francisco. Funding sources did not have a direct role in the design, analysis, or approval of this manuscript.

## Consortium Appendix A, IMPACC Network

Elaine F. Reed, Joanna Schaenman, Ramin Salehi-rad, Adreanne M. Rivera, Harry Pickering, Subha Sen, David Elashoff, Dawn C. Ward, Jenny Brook, Estefania Ramires-Sanchez, Megan Llamas, Claudia Perdomo, Clara E. Magyar, Jennifer Fulcher, David J. Erle, Carolyn S. Calfee, Carolyn M. Hendrickson, Kirsten N. Kangelaris, Viet Nguyen, Deanna Lee, Suzanna Chak, Rajani Ghale, Ana Gonzalez, Alejandra Jauregui, Carolyn Leroux, Luz Torres Altamirano, Ahmad Sadeed Rashid, Andrew Willmore, Prescott G. Woodruff, Matthew F. Krummel, Sidney Carrillo, Alyssa Ward, Charles R. Langelier, Ravi Patel, Michael Wilson, Ravi Dandekar, Bonny Alvarenga, Jayant Rajan, Walter Eckalbar, Andrew W. Schroeder, Gabriela K. Fragiadakis, Alexandra Tsitsiklis, Eran Mick, Yanedth Sanchez Guerrero, Christina Love, Lenka Maliskova, Michael Adkisson, Aleksandra Leligdowicz, Alexander Beagle, Arjun Rao, Austin Sigman, Bushra Samad, Cindy Curiel, Cole Shaw, Gayelan Tietje-Ulrich, Jeff Milush, Jonathan Singer, Joshua J. Vasquez, Kevin Tang, Legna Betancourt, Lekshmi Santhosh, Logan Pierce, Maria Tecero Paz, Michael M. Matthay, Neeta Thakur, Nicklaus Rodriguez, Nicole Sutter, Norman Jones, Pratik Sinha, Priya Prasad, Raphael Lota, Sadeed Rashid, Saurabh Asthana, Sharvari Bhide, Tasha Lea, Yumiko Abe-Jones, Lauren I. R. Ehrlich, Esther Melamed, Cole Maguire, Dennis Wylie, Justin F. Rousseau, Kerin C. Hurley, Janelle N. Geltman, Nadia Siles, Jacob E. Rogers, Pablo Guaman Tipan, Nadine Rouphael, Steven E. Bosinger, Arun K. Boddapati, Greg K. Tharp, Kathryn L. Pellegrini, Brandi Johnson, Bernadine Panganiban, Christopher Huerta, Evan J. Anderson, Hady Samaha, Jonathan E. Sevransky, Laurel Bristow, Elizabeth Beagle, David Cowan, Sydney Hamilton, Thomas Hodder, Amer Bechnak, Andrew Cheng, Aneesh Mehta, Caroline R. Ciric, Christine Spainhour, Erin Carter, Erin M. Scherer, Jacob Usher, Kieffer Hellmeister, Laila Hussaini, Lauren Hewitt, Nina Mcnair, Susan Pereira Ribeiro, Sonia Wimalasena, Jordan P. Metcalf, Nelson I. Agudelo Higuita, Lauren A. Sinko, J. Leland Booth, Douglas A. Drevets, Brent R. Brown, Mark A. Atkinson, Scott C. Brakenridge, Ricardo F. Ungaro, Brittany Roth Manning, Lyle Moldawer, William B. Messer, Catherine L. Hough, Sarah A. R. Siegel, Peter E. Sullivan, Zhengchun Lu, Amanda E. Brunton, Matthew Strand, Zoe L. Lyski, Felicity J. Coulter, Courtney Micheletti, Matthew C. Altman, Naresh Doni Jayavelu, Scott Presnell, Bernard Kohr, Tomasz Jancsyk, Azlann Arnett, Patrice M. Becker, Alison D. Augustine, Steven M. Holland, Lindsey B. Rosen, Serena Lee, Tatyana Vaysman, Al Ozonoff, Joann Diray-Arce, Jing Chen, Alvin T. Kho, Carly E. Milliren, Annmarie Hoch, Ana C. Chang, Kerry McEnaney, Caitlin Syphurs, Brenda Barton, Claudia Lentucci, Maimouna D. Murphy, Mehmet Saluvan, Tanzia Shaheen, Shanshan Liu, Marisa Albert, Arash Nemati Hayati, Robert Bryant, James Abraham, Mitchell Cooney, Meagan Karoly, Ofer Levy, Hanno Steen, Patrick van Zalm, Benoit Fatou, Kinga K. Smolen, Arthur Viode, Simon van Haren, Meenakshi Jha, David Stevenson, Sanya Thomas, Boryana Petrova, Naama Kanarek, Ana Fernandez-Sesma, Viviana Simon, Florian Krammer, Harm Van Bakel, Seunghee Kim-Schulze, Ana Silvia Gonzalez-Reiche, Jingjing Qi, Brian Lee, Juan Manuel Carreño, Gagandeep Singh, Ariel Raskin, Johnstone Tcheou, Zain Khalil, Adriana van de Guchte, Keith Farrugia, Zenab Khan, Geoffrey Kelly, Komal Srivastava, Lily Q. Eaker, Maria C. Bermúdez-González, Lubbertus C. F. Mulder, Katherine F. Beach, Miti Saksena, Deena Altman, Erna Kojic, Levy A. Sominsky, Arman Azad, Dominika Bielak, Hisaaki Kawabata, Temima Yellin, Miriam Fried, Leeba Sullivan, Sara Morris, Giulio Kleiner, Daniel Stadlbauer, Jayeeta Dutta, Hui Xie, Manishkumar Patel, Kai Nie, Brian Monahan, David A. Hafler, Ruth R. Montgomery, Albert C. Shaw, Steven H. Kleinstein, Jeremy P. Gygi, Dylan Duchen, Shrikant Pawar, Anna Konstorum, Ernie Chen, Chris Cotsapas, Xiaomei Wang, Charles Dela Cruz, Akiko Iwasaki, Subhasis Mohanty, Allison Nelson, Yujiao Zhao, Shelli Farhadian, Hiromitsu Asashima, Omkar Chaudhary, Andreas Coppi, John Fournier, M. Catherine Muenker, Khadir Raddassi, Michael Rainone, William Ruff, Syim Salahuddin, Wade L. Shulz, Pavithra Vijayakumar, Haowei Wang, Esio Wunder Jr., H. Patrick Young, Albert I. Ko, Gisela Gabernet, Denise Esserman, Leying Guan, Anderson Brito, Jessica Rothman, Nathan D. Grubaugh, Kexin Wang, Leqi Xu, Holden Maecker, Bali Pulendran, Kari C. Nadeau, Yael Rosenberg-Hasson, Michael Leipold, Natalia Sigal, Angela Rogers, Andrea Fernandes, Monali Manohar, Evan Do, Iris Chang, Alexandra S. Lee, Catherine Blish, Henna Naz Din, Jonasel Roque, Linda N. Geng, Maja Artandi, Mark M. Davis, Neera Ahuja, Samuel S. Yang, Sharon Chinthrajah, Thomas Hagan, Tyson H. Holmes, Koji Abe, Lindsey R. Baden, Kevin Mendez, Jessica Lasky-Su, Alexandra Tong, Rebecca Rooks, Michael Desjardins, Amy C. Sherman, Stephen R. Walsh, Xhoi Mitre, Jessica Cauley, Xiaofang Li, Bethany Evans, Christina Montesano, Jose Humberto Licona, Jonathan Krauss, Nicholas C. Issa, Jun Bai Park Chang, Natalie Izaguirre, David B. Corry, Farrah Kheradmand, Li-Zhen Song, Ebony Nelson, Monica Kraft, Chris Bime, Jarrod Mosier, Heidi Erickson, Ron Schunk, Hiroki Kimura, Michelle Conway, Dave Francisco, Allyson Molzahn, Connie Cathleen Wilson, Ron Schunk, Trina Hughes, Bianca Sierra, Jordan Oberhaus, Faheem W. Guirgis, Brittney Borresen, Matthew L. Anderson, Bjoern Peters, James A. Overton, Randi Vita, Kerstin Westendorf, Scott J. Tebbutt, Casey P. Shannon, Rafick-Pierre Sekaly, Slim Fourati, Grace A. McComsey, Paul Harris, Scott Sieg, George Yendewa, Mary Consolo, Heather Tribout, Susan Pereira Ribeiro, Charles B. Cairns, Elias K. Haddad, Michele A. Kutzler, Mariana Bernui, Gina Cusimano, Jennifer Connors, Kyra Woloszczuk, David Joyner, Carolyn Edwards, Edward Lee, Edward Lin, Nataliya Melnyk, Debra L. Powell, James N. Kim, I. Michael Goonewardene, Brent Simmons, Cecilia M. Smith, Mark Martens, Brett Croen, Nicholas C. Semenza, Mathew R. Bell, Sara Furukawa, Renee McLin, George P. Tegos, Brandon Rogowski, Nathan Mege, Kristen Ulring, Pam Schearer, Judie Sheidy, Crystal Nagle, James A. Overton, Scott R. Hutton, Greg Michelotti, Kari Wong, Adeeb Rahman & Vicki Seyfert-Margolis.

## Consortium Appendix B, COMET Consortium

K. Mark Ansel, Stephanie Christenson, Michael Adkisson, Walter Eckalbar, Lenka Maliskova, Andrew Schroeder, Raymund Bueno, Gracie Gordon, George Hartoularos, Divya Kushnoor, David Lee, Elizabeth McCarthy, Anton Ogorodnikov, Matthew Spitzer, Kamir Hiam, Yun S. Song, Yang Sun, Erden Tumurbaatar, Monique van der Wijst, Alexander Whatley, Chayse Jones, Saharai Caldera, Catherine DeVoe, Paula Hayakawa Serpa, Christina Love, Eran Mick, Maira Phelps, Alexandra Tsitsiklis, Carolyn Leroux, Sadeed Rashid, Nicklaus Rodriguez, Kevin Tang, Luz Torres Altamirano, Aleksandra Leligdowicz, Michael Matthay, Michael Wilson, Jimmie Ye, Suzanna Chak, Rajani Ghale, Alejandra Jauregui, Deanna Lee, Viet Nguyen, Austin Sigman, Kirsten N. Kangelaris, Saurabh Asthana, Zachary Collins, Ravi Patel, Arjun Rao, Bushra Samad, Cole Shaw, Andrew Willmore, Tasha Lea, Gabriela K. Fragiadakis, Carolyn S. Calfee, David J. Erle, Carolyn M. Hendrickson, Matthew F. Krummel, Charles R. Langelier, Prescott G. Woodruff, Sidney C. Haller, Alyssa Ward, Norman Jones, Jeff Milush, Vincent Chan, Nayvin Chew, Alexis Combes, Tristan Courau, Kenneth Hu, Billy Huang, Nitasha Kumar, Salman Mahboob, Priscila Muñoz-Sandoval, Randy Parada, Gabriella Reeder, Alan Shen, Jessica Tsui, Shoshana Zha & Wandi S. Zhu.

## Consortium Appendix C, EARLI Consortium

Narges Alipanah-Lechner, Kim Bardillon, Carolyn S. Calfee, Suzanna Chak, Olivia Chao Sidney A. Carrillo, Taarini Hariharan, Carolyn M. Hendrickson, Charles R Langelier, Deanna Lee, Carolyn Leroux, Chelsea Lin, Michael A. Matthay, Lucile P. A. Neyton, Angelika Ringor, Aartik Sarma, Emma Schmiege, Natasha Spottiswoode, Kathryn M. Sullivan, Melanie F Weingart, Andrew Willmore, Hanjing Zhuo.

## Conflict of interest statement

The Icahn School of Medicine at Mount Sinai has filed patent applications relating to SARS-CoV-2 serological assays, NDV-based SARS-CoV-2 vaccines influenza virus vaccines and influenza virus therapeutics which list Florian Krammer as co-inventor and he has received royalty payments from some of these patents. Mount Sinai has spun out a company, Kantaro, to market serological tests for SARS-CoV-2 and another company, Castlevax, to develop SARS-CoV-2 vaccines, and Florian Krammer is co-founder and scientific advisory board member. Florian Krammer has consulted for Merck, GSK, Sanofi, Curevac, Seqirus and Pfizer and is currently consulting for 3rd Rock Ventures, Gritstone and Avimex, as well as collaborating with Dynavax on influenza vaccine development and with VIR on influenza virus therapeutics. Ofer Levy is a named inventor on patents held by Boston Children’s Hospital relating to vaccine adjuvants and human in vitro platforms that model vaccine action. His laboratory has received research support from GlaxoSmithKline (GSK) and is a co-founder of and advisor to Ovax, Inc. Charles Cairns serves as a consultant to bioMerieux and is funded for a grant from Bill & Melinda Gates Foundation. James A Overton is a consultant at Knocean Inc. Jessica Lasky-Su serves as a scientific advisor of Precion Inc. Scott R. Hutton, Greg Michelloti and Kari Wong are employees of Metabolon Inc. Vicki Seyfer-Margolis is a current employee of MyOwnMed. Nadine Rouphael reports grants or contracts with Merck, Sanofi, Pfizer, Vaccine Company, Quidel, Lilly and Immorna, and has participated on data safety monitoring boards for Moderna, Sanofi, Seqirus, Pfizer, EMMES, ICON, BARDA, Imunon, CyanVac and Micron. Nadine Rouphael has also received support for meetings/travel from Sanofi and Moderna and honoraria from Virology Education. Adeeb Rahman is a current employee of Immunai Inc. Steven Kleinstein is a consultant related to ImmPort data repository for Peraton. Nathan Grabaugh is a consultant for Tempus Labs and the National Basketball Association. Akiko Iwasaki is a consultant for 4BIO, Blue Willow Biologics, Revelar Biotherapeutics, RIGImmune, Xanadu Bio, Paratus Sciences. Monika Kraft receives research funds paid to her institution from NIH, ALA; Sanofi, Astra-Zeneca for work in asthma, serves as a consultant for Astra-Zeneca, Sanofi, Chiesi, GSK for severe asthma; is a co-founder and CMO for RaeSedo, Inc, a company created to develop peptidomimetics for treatment of inflammatory lung disease. Esther Melamed received research funding from Babson Diagnostics and honorarium from Multiple Sclerosis Association of America and has served on the advisory boards of Genentech, Horizon, Teva, and Viela Bio. Carolyn Calfee receives research funding from NIH, FDA, DOD, Roche-Genentech and Quantum Leap Healthcare Collaborative as well as consulting services for Janssen, Vasomune, Gen1e Life Sciences, NGMBio, and Cellenkos. Wade Schulz was an investigator for a research agreement, through Yale University, from the Shenzhen Center for Health Information for work to advance intelligent disease prevention and health promotion; collaborates with the National Center for Cardiovascular Diseases in Beijing; is a technical consultant to Hugo Health, a personal health information platform; cofounder of Refactor Health, an AI-augmented data management platform for health care; and has received grants from Merck and Regeneron Pharmaceutical for research related to COVID-19. Grace A McComsey received research grants from Redhill, Cognivue, Pfizer, and Genentech, and served as a research consultant for Gilead, Merck, Viiv/GSK, and Jenssen. Linda N. Geng received research funding paid to her institution from Pfizer, Inc.

## Notes

### Competing Interest Statement

Please see manuscript for full description.

### Author Declarations

The Department of Health and Human Services Office for Human Research Protections determined that the IMPACC study protocol met criteria for a public health surveillance exception [45CFR46.102(l)(2)], and the study was approved by each institutional review board (IRB) through this exception (12 sites) or by pre-approved biobanking protocols (3 sites).

## REFERENCES

1. Flisiak R, Rzymski P, Zarębska-Michaluk D, et al. Variability in the Clinical Course of COVID-19 in a Retrospective Analysis of a Large Real-World Database. Viruses. 2023;15(1):149. doi:10.3390/v15010149

2. Tenforde MW, Billig Rose E, Lindsell CJ, et al. Characteristics of Adult Outpatients and Inpatients with COVID-19 - 11 Academic Medical Centers, United States, March-May 2020. MMWR Morb Mortal Wkly Rep. 2020;69(26):841-846. doi:10.15585/mmwr.mm6926e3

3. Funada S, Yoshioka T, Luo Y, et al. Global Trends in Highly Cited Studies in COVID-19 Research. JAMA Network Open. 2023;6(9):e2332802. doi:10.1001/jamanetworkopen.2023.32802

4. Arunachalam PS, Wimmers F, Mok CKP, et al. Systems biological assessment of immunity to mild versus severe COVID-19 infection in humans. Science. 2020;369(6508):1210-1220. doi:10.1126/science.abc6261

5. Gygi JP, Maguire C, Patel RK, et al. Integrated longitudinal multiomics study identifies immune programs associated with acute COVID-19 severity and mortality. J Clin Invest. 2024;134(9). doi:10.1172/JCI176640

6. Diray-Arce J, Fourati S, Doni Jayavelu N, et al. Multi-omic longitudinal study reveals immune correlates of clinical course among hospitalized COVID-19 patients. Cell Reports Medicine. 2023;4(6):101079. doi:10.1016/j.xcrm.2023.101079

7. Emanuel EJ, Persad G, Upshur R, et al. Fair Allocation of Scarce Medical Resources in the Time of Covid-19. N Engl J Med. 2020;382(21):2049–2055. doi:10.1056/NEJMsb2005114

8. Vergano M, Bertolini G, Giannini A, et al. Clinical ethics recommendations for the allocation of intensive care treatments in exceptional, resource-limited circumstances: the Italian perspective during the COVID-19 epidemic. Crit Care. 2020;24(1):165. doi:10.1186/s13054-020-02891-w

9. Viele K, Girard TD. Risk, Results, and Costs: Optimizing Clinical Trial Efficiency through Prognostic Enrichment. Am J Respir Crit Care Med. 2021;203(6):671–672. doi:10.1164/rccm.202009-3649ED

10. Stanski NL, Wong HR. Prognostic and predictive enrichment in sepsis. Nat Rev Nephrol. 2020;16(1):20–31. doi:10.1038/s41581-019-0199-3

11. Romero Starke K, Reissig D, Petereit-Haack G, Schmauder S, Nienhaus A, Seidler A. The isolated effect of age on the risk of COVID-19 severe outcomes: a systematic review with meta-analysis. BMJ Glob Health. 2021;6(12):e006434. doi:10.1136/bmjgh-2021-006434

12. Zhang X, Tan Y, Ling Y, et al. Viral and host factors related to the clinical outcome of COVID-19. Nature. 2020;583(7816):437-440. doi:10.1038/s41586-020-2355-0

13. Brodin P. Immune determinants of COVID-19 disease presentation and severity. Nat Med. 2021;27(1):28–33. doi:10.1038/s41591-020-01202-8

14. Sumi T, Harada K. Immune response to SARS-CoV-2 in severe disease and long COVID-19. iScience. 2022;25(8):104723. doi:10.1016/j.isci.2022.104723

15. Malik P, Patel U, Mehta D, et al. Biomarkers and outcomes of COVID-19 hospitalisations: systematic review and meta-analysis. BMJ Evid Based Med. 2021;26(3):107–108. doi:10.1136/bmjebm-2020-111536

16. Ponti G, Maccaferri M, Ruini C, Tomasi A, Ozben T. Biomarkers associated with COVID-19 disease progression. Crit Rev Clin Lab Sci. Published online June 5, 2020:1–11. doi:10.1080/10408363.2020.1770685

17. Wang M, Wu D, Liu CH, et al. Predicting progression to severe COVID-19 using the PAINT score. BMC Infectious Diseases. 2022;22(1):498. doi:10.1186/s12879-022-07466-4

18. Zhao Z, Chen A, Hou W, et al. Prediction model and risk scores of ICU admission and mortality in COVID-19. PLoS One. 2020;15(7):e0236618. doi:10.1371/journal.pone.0236618

19. Raman G, Ashraf B, Demir YK, et al. Machine learning prediction for COVID-19 disease severity at hospital admission. BMC Med Inform Decis Mak. 2023;23(1):46. doi:10.1186/s12911-023-02132-4

20. Liang W, Liang H, Ou L, et al. Development and Validation of a Clinical Risk Score to Predict the Occurrence of Critical Illness in Hospitalized Patients With COVID-19. JAMA Intern Med. 2020;180(8):1081–1089. doi:10.1001/jamainternmed.2020.2033

21. Buttia C, Llanaj E, Raeisi-Dehkordi H, et al. Prognostic models in COVID-19 infection that predict severity: a systematic review. Eur J Epidemiol. 2023;38(4):355–372. doi:10.1007/s10654-023-00973-x

22. Chaussabel D, Pascual V, Banchereau J. Assessing the human immune system through blood transcriptomics. BMC Biol. 2010;8:84. doi:10.1186/1741-7007-8-84

23. Burel JG, Babor M, Pomaznoy M, et al. Host Transcriptomics as a Tool to Identify Diagnostic and Mechanistic Immune Signatures of Tuberculosis. Front Immunol. 2019;10:221. doi:10.3389/fimmu.2019.00221

24. Tsalik EL, Henao R, Nichols M, et al. Host gene expression classifiers diagnose acute respiratory illness etiology. Sci Transl Med. 2016;8(322):322ra11. doi:10.1126/scitranslmed.aad6873

25. Langelier C, Kalantar KL, Moazed F, et al. Integrating host response and unbiased microbe detection for lower respiratory tract infection diagnosis in critically ill adults. Proc Natl Acad Sci U S A. 2018;115(52):E12353–E12362. doi:10.1073/pnas.1809700115

26. Mick E, Tsitsiklis A, Kamm J, et al. Integrated host/microbe metagenomics enables accurate lower respiratory tract infection diagnosis in critically ill children. J Clin Invest. 2023;133(7). doi:10.1172/JCI165904

27. Lydon EC, Ko ER, Tsalik EL. The host response as a tool for infectious disease diagnosis and management. Expert Review of Molecular Diagnostics. 2018;18(8):723–738. doi:10.1080/14737159.2018.1493378

28. Buturovic L, Zheng H, Tang B, et al. A 6-mRNA host response classifier in whole blood predicts outcomes in COVID-19 and other acute viral infections. Sci Rep. 2022;12(1):889. doi:10.1038/s41598-021-04509-9

29. Daenen K, Tong-Minh K, Liesenfeld O, et al. A Transcriptomic Severity Classifier IMX-SEV-3b to Predict Mortality in Intensive Care Unit Patients with COVID-19: A Prospective Observational Pilot Study. J Clin Med. 2023;12(19):6197. doi:10.3390/jcm12196197

30. Baghela A, An A, Zhang P, et al. Predicting severity in COVID-19 disease using sepsis blood gene expression signatures. Sci Rep. 2023;13(1):1247. doi:10.1038/s41598-023-28259-y

31. Armignacco R, Carlier N, Jouinot A, et al. Whole blood transcriptome signature predicts severe forms of COVID-19: Results from the COVIDeF cohort study. Funct Integr Genomics. 2024;24(3):1–14. doi:10.1007/s10142-024-01359-2

32. Kangelaris KN, Prakash A, Liu KD, et al. Increased expression of neutrophil-related genes in patients with early sepsis-induced ARDS. Am J Physiol Lung Cell Mol Physiol. 2015;308(11):L1102–1113. doi:10.1152/ajplung.00380.2014

33. Maddali MV, Churpek M, Pham T, et al. Validation and Utility of ARDS Subphenotypes Identified by Machine Learning Models Using Clinical Data: An Observational Multi-Cohort Retrospective Analysis. Lancet Respir Med. 2022;10(4):367–377. doi:10.1016/S2213-2600(21)00461-6

34. Krzyzak L, Seitz C, Urbat A, et al. CD83 Modulates B Cell Activation and Germinal Center Responses. The Journal of Immunology. 2016;196(9):3581–3594. doi:10.4049/jimmunol.1502163

35. Aerts-Toegaert C, Heirman C, Tuyaerts S, et al. CD83 expression on dendritic cells and T cells: correlation with effective immune responses. Eur J Immunol. 2007;37(3):686–695. doi:10.1002/eji.200636535

36. Orlov SN, Thorin-Trescases N, Pchejetski D, et al. Na+/K+ pump and endothelial cell survival: [Na+]i/[K+]i-independent necrosis triggered by ouabain, and protection against apoptosis mediated by elevation of [Na+]i. Pflugers Arch. 2004;448(3):335–345. doi:10.1007/s00424-004-1262-9

37. de Alwis N, Beard S, Binder NK, et al. DAAM2 is elevated in the circulation and placenta in pregnancies complicated by fetal growth restriction and is regulated by hypoxia. Sci Rep. 2021;11(1):5540. doi:10.1038/s41598-021-84785-7

38. Jia X, Crawford JC, Gebregzabher D, et al. High expression of oleoyl-ACP hydrolase underpins life-threatening respiratory viral diseases. Cell. 2024;187(17):4586–4604.e20. doi:10.1016/j.cell.2024.07.026

39. Neyton LPA, Patel RK, Sarma A, et al. Distinct pulmonary and systemic effects of dexamethasone in severe COVID-19. Nat Commun. 2024;15(1):5483. doi:10.1038/s41467-024-49756-2

40. Wang D, Hu B, Hu C, et al. Clinical Characteristics of 138 Hospitalized Patients With 2019 Novel Coronavirus-Infected Pneumonia in Wuhan, China. JAMA. 2020;323(11):1061–1069. doi:10.1001/jama.2020.1585

41. Bertagnolio S, Thwin SS, Silva R, et al. Clinical features of, and risk factors for, severe or fatal COVID-19 among people living with HIV admitted to hospital: analysis of data from the WHO Global Clinical Platform of COVID-19. The Lancet HIV. 2022;9(7):e486–e495. doi:10.1016/S2352-3018(22)00097-2

42. Kim L, Garg S, O’Halloran A, et al. Risk Factors for Intensive Care Unit Admission and In-hospital Mortality Among Hospitalized Adults Identified through the US Coronavirus Disease 2019 (COVID-19)-Associated Hospitalization Surveillance Network (COVID-NET). Clin Infect Dis. 2021;72(9):e206–e214. doi:10.1093/cid/ciaa1012

43. Amrute JM, Perry AM, Anand G, et al. Cell specific peripheral immune responses predict survival in critical COVID-19 patients. Nat Commun. 2022;13(1):882. doi:10.1038/s41467-022-28505-3

44. World Health Organization. WHO COVID-19 Dashboard. Accessed February 7, 2025. https://data.who.int/dashboards/covid19/cases

45. Gliddon HD, Herberg JA, Levin M, Kaforou M. Genome-wide host RNA signatures of infectious diseases: discovery and clinical translation. Immunology. 2018;153(2):171–178. doi:10.1111/imm.12841

46. Perez-Pons M, Molinero M, Benítez ID, et al. MicroRNA-centered theranostics for pulmoprotection in critical COVID-19. Mol Ther Nucleic Acids. 2024;35(1):102118. doi:10.1016/j.omtn.2024.102118

47. Singh MS, Arun PPS, Ansari MA, Singh MS, Arun PPS, Ansari MA. Unveiling common markers in COVID-19: ADAMTS2, PCSK9, and OLAH emerged as key differential gene expression profiles in PBMCs across diverse disease conditions. AIMSMOLES. 2024;11(2):189-205. doi:10.3934/molsci.2024011

48. Venet M, Ribeiro MS, Décembre E, et al. Severe COVID-19 patients have impaired plasmacytoid dendritic cell-mediated control of SARS-CoV-2. Nat Commun. 2023;14(1):694. doi:10.1038/s41467-023-36140-9

49. Riaz B, Islam SMS, Ryu HM, Sohn S. CD83 Regulates the Immune Responses in Inflammatory Disorders. Int J Mol Sci. 2023;24(3):2831. doi:10.3390/ijms24032831

50. Buckley JF, Singer M, Clapp LH. Role of KATP channels in sepsis. Cardiovascular Research. 2006;72(2):220–230. doi:10.1016/j.cardiores.2006.07.011

51. Kryvenko V, Vadász I. Molecular mechanisms of Na,K-ATPase dysregulation driving alveolar epithelial barrier failure in severe COVID-19. Am J Physiol Lung Cell Mol Physiol. 2021;320(6):L1186-L1193. doi:10.1152/ajplung.00056.2021

52. Wang CY, Zuo Z, Jo J, et al. Daam2 phosphorylation by CK2α negatively regulates Wnt activity during white matter development and injury. Proc Natl Acad Sci U S A. 2023;120(35):e2304112120. doi:10.1073/pnas.2304112120

53. Pereira CP, Bachli EB, Schoedon G. The wnt pathway: a macrophage effector molecule that triggers inflammation. Curr Atheroscler Rep. 2009;11(3):236–242. doi:10.1007/s11883-009-0036-4

54. Olsen JJ, Pohl SÖG, Deshmukh A, et al. The Role of Wnt Signalling in Angiogenesis. Clin Biochem Rev. 2017;38(3):131–142.

55. Yue J, Mo L, Zeng G, et al. Inhibition of neutrophil extracellular traps alleviates blood-brain barrier disruption and cognitive dysfunction via Wnt3/β-catenin/TCF4 signaling in sepsis-associated encephalopathy. J Neuroinflammation. 2025;22(1):87. doi:10.1186/s12974-025-03395-6

56. Lee HK, Chaboub LS, Zhu W, et al. Daam2-PIP5K Is a Regulatory Pathway for Wnt Signaling and Therapeutic Target for Remyelination in the CNS. Neuron. 2015;85(6):1227–1243. doi:10.1016/j.neuron.2015.02.024

57. Nakaya M aki, Gudmundsson KO, Komiya Y, et al. Placental defects lead to embryonic lethality in mice lacking the Formin and PCP proteins Daam1 and Daam2. PLOS ONE. 2020;15(4):e0232025. doi:10.1371/journal.pone.0232025

58. Lui KO, Ma Z, Dimmeler S. SARS-CoV-2 induced vascular endothelial dysfunction: direct or indirect effects? Cardiovascular Research. 2024;120(1):34–43. doi:10.1093/cvr/cvad191

59. Merad M, Blish CA, Sallusto F, Iwasaki A. The immunology and immunopathology of COVID-19. Science. 2022;375(6585):1122-1127. doi:10.1126/science.abm8108

60. Bost P, De Sanctis F, Canè S, et al. Deciphering the state of immune silence in fatal COVID-19 patients. Nat Commun. 2021;12:1428. doi:10.1038/s41467-021-21702-6

61. Hadjadj J, Yatim N, Barnabei L, et al. Impaired type I interferon activity and inflammatory responses in severe COVID-19 patients. Science. 2020;369(6504):718-724. doi:10.1126/science.abc6027

62. Middleton EA, He XY, Denorme F, et al. Neutrophil extracellular traps contribute to immunothrombosis in COVID-19 acute respiratory distress syndrome. Blood. 2020;136(10):1169–1179. doi:10.1182/blood.2020007008

63. Del Valle DM, Kim-Schulze S, Huang HH, et al. An inflammatory cytokine signature predicts COVID-19 severity and survival. Nat Med. 2020;26(10):1636–1643. doi:10.1038/s41591-020-1051-9

64. Lucas C, Wong P, Klein J, et al. Longitudinal analyses reveal immunological misfiring in severe COVID-19. Nature. 2020;584(7821):463-469. doi:10.1038/s41586-020-2588-y

65. A blood atlas of COVID-19 defines hallmarks of disease severity and specificity. Cell. 2022;185(5):916-938.e58. doi:10.1016/j.cell.2022.01.012

66. Varga Z, Flammer AJ, Steiger P, et al. Endothelial cell infection and endotheliitis in COVID-19. The Lancet. 2020;395(10234):1417–1418. doi:10.1016/S0140-6736(20)30937-5

67. Hopp MT, Domingo-Fernández D, Gadiya Y, et al. Linking COVID-19 and Heme-Driven Pathophysiologies: A Combined Computational–Experimental Approach. Biomolecules. 2021;11(5):644. doi:10.3390/biom11050644

68. Mendonça MM, da Cruz KR, Pinheiro D da S, et al. Dysregulation in erythrocyte dynamics caused by SARS-CoV-2 infection: possible role in shuffling the homeostatic puzzle during COVID-19. Hematology, Transfusion and Cell Therapy. 2022;44(2):235-245. doi:10.1016/j.htct.2022.01.005

69. Gioia U, Tavella S, Martínez-Orellana P, et al. SARS-CoV-2 infection induces DNA damage, through CHK1 degradation and impaired 53BP1 recruitment, and cellular senescence. Nat Cell Biol. 2023;25(4):550–564. doi:10.1038/s41556-023-01096-x

70. Grand RJ. SARS-CoV-2 and the DNA damage response. J Gen Virol. 2023;104(11):001918. doi:10.1099/jgv.0.001918

71. Carlini V, Noonan DM, Abdalalem E, et al. The multifaceted nature of IL-10: regulation, role in immunological homeostasis and its relevance to cancer, COVID-19 and post-COVID conditions. Front Immunol. 2023;14:1161067. doi:10.3389/fimmu.2023.1161067

72. Lu L, Zhang H, Dauphars DJ, He YW. A Potential Role of Interleukin 10 in COVID-19 Pathogenesis. Trends Immunol. 2021;42(1):3–5. doi:10.1016/j.it.2020.10.012

73. Beltrán-García J, Osca-Verdegal R, Pallardó FV, et al. Sepsis and Coronavirus Disease 2019: Common Features and Anti-Inflammatory Therapeutic Approaches. Crit Care Med. 2020;48(12):1841-1844. doi:10.1097/CCM.0000000000004625

74. Olwal CO, Nganyewo NN, Tapela K, et al. Parallels in Sepsis and COVID-19 Conditions: Implications for Managing Severe COVID-19. Front Immunol. 2021;12:602848. doi:10.3389/fimmu.2021.602848

75. An AY, Baghela A, Zhang P, et al. Severe COVID-19 and non-COVID-19 severe sepsis converge transcriptionally after a week in the intensive care unit, indicating common disease mechanisms. Front Immunol. 2023;14:1167917. doi:10.3389/fimmu.2023.1167917

76. Ozonoff A, Schaenman J, Jayavelu ND, et al. Phenotypes of disease severity in a cohort of hospitalized COVID-19 patients: Results from the IMPACC study. EBioMedicine. 2022;83:104208. doi:10.1016/j.ebiom.2022.104208

77. IMPACC Manuscript Writing Team, IMPACC Network Steering Committee. Immunophenotyping assessment in a COVID-19 cohort (IMPACC): A prospective longitudinal study. Sci Immunol. 2021;6(62):eabf3733. doi:10.1126/sciimmunol.abf3733

78. Kalantar KL, Neyton L, Abdelghany M, et al. Integrated host-microbe plasma metagenomics for sepsis diagnosis in a prospective cohort of critically ill adults. Nat Microbiol. 2022;7(11):1805–1816. doi:10.1038/s41564-022-01237-2

79. Maechler M, Todorov V, Ruckstuhl A, Salibian-Barrera M, Koller M, Conceicao ELT. robustbase: Basic Robust Statistics. Published online September 27, 2024:0.99–4-1. doi:10.32614/CRAN.package.robustbase

80. Ritchie ME, Phipson B, Wu D, et al. limma powers differential expression analyses for RNA-sequencing and microarray studies. Nucleic Acids Res. 2015;43(7):e47. doi:10.1093/nar/gkv007

81. Law CW, Chen Y, Shi W, Smyth GK. voom: Precision weights unlock linear model analysis tools for RNA-seq read counts. Genome Biol. 2014;15(2):R29. doi:10.1186/gb-2014-15-2-r29

82. Korotkevich G, Sukhov V, Budin N, Shpak B, Artyomov MN, Sergushichev A. Fast gene set enrichment analysis. Published online February 1, 2021:060012. doi:10.1101/060012

83. Sidiropoulos K, Viteri G, Sevilla C, et al. Reactome enhanced pathway visualization. Bioinformatics. 2017;33(21):3461–3467. doi:10.1093/bioinformatics/btx441

84. Love MI, Huber W, Anders S. Moderated estimation of fold change and dispersion for RNA-seq data with DESeq2. Genome Biology. 2014;15(12):550. doi:10.1186/s13059-014-0550-8

85. Kuhn M. Building Predictive Models in R Using the caret Package. Journal of Statistical Software. 2008;28:1–26. doi:10.18637/jss.v028.i05

86. Tibshirani R. Regression Shrinkage and Selection via the Lasso. Journal of the Royal Statistical Society Series B (Methodological). 1996;58(1):267–288.

87. Friedman J, Hastie T, Tibshirani R, et al. glmnet: Lasso and Elastic-Net Regularized Generalized Linear Models. Published online August 22, 2023. Accessed March 17, 2025. https://cran.r-project.org/web/packages/glmnet/index.html

88. Robin X, Turck N, Hainard A, et al. pROC: an open-source package for R and S+ to analyze and compare ROC curves. BMC Bioinformatics. 2011;12(1):77. doi:10.1186/1471-2105-12-77

