## Supplementary Materials for "Minimalistic Transcriptomic Signatures Permit Accurate Early Prediction of COVID-19 Mortality"

1. **SUPPLEMENTARY FIGURES**
2. **SUPPLEMENTARY TABLES**
3. **SUPPLEMENTARY DATA FILES**

### SUPPLEMENTARY FIGURES

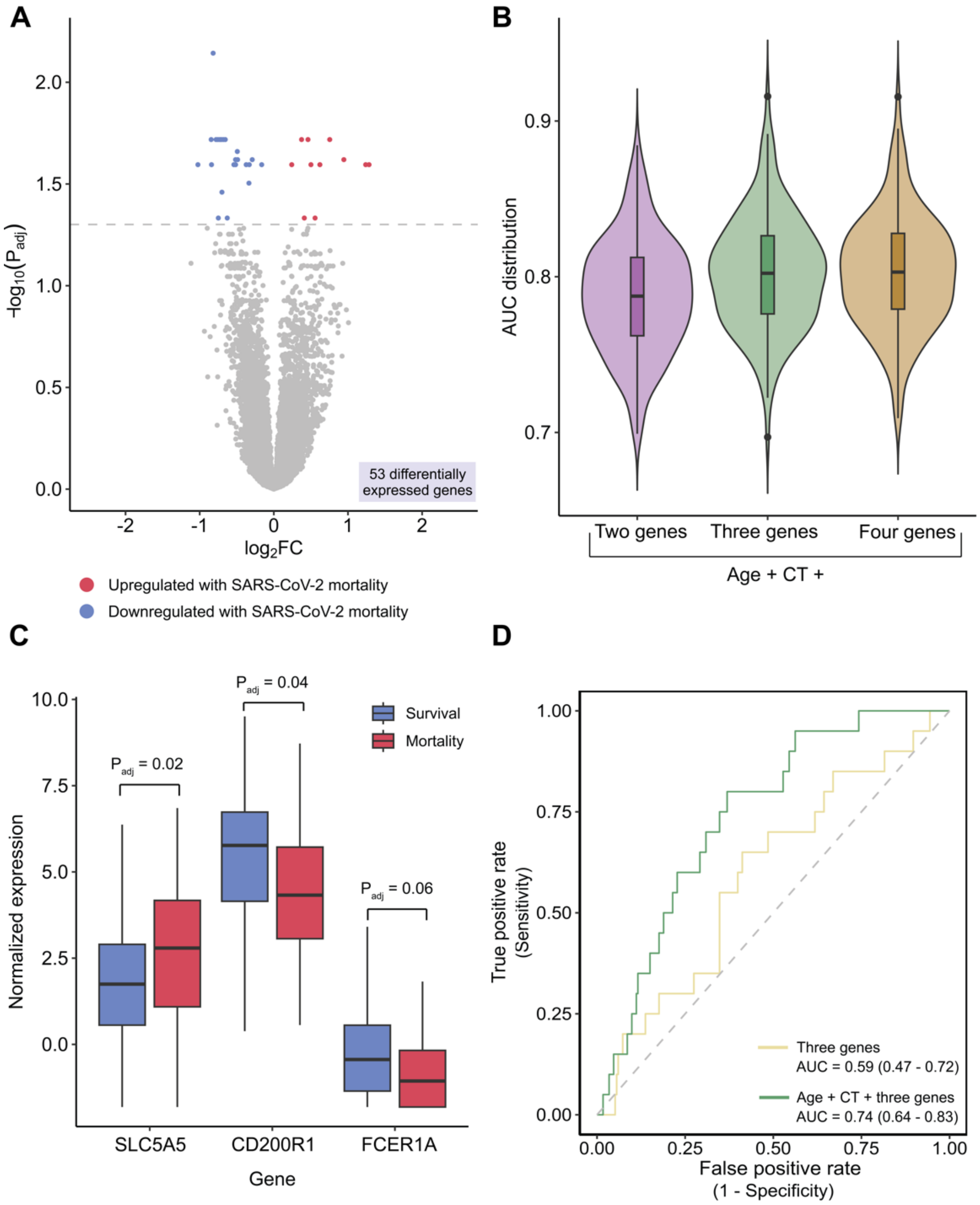


**Figure S1. Upper respiratory tract differential expression and host-viral classifier development and evaluation. A** Volcano plot demonstrating the 53 differentially expressed genes between mortality and survival in the upper respiratory tract in the IMPACC cohort (red = upregulated with mortality, blue = downregulated with mortality), using a Benjamini-Hochberg adjusted p-value of 0.05. **B.** Violin plots showing the area under the curve (AUC) distribution for each of the upper respiratory tract classifiers, evaluated in the training cohort. **C.** Boxplots comparing the $\log_2$ counts per million normalized gene expression of the three-gene classifier genes *(SLC5A5, CD200R1*, *FCER1A*) between survival (blue) and mortality (red) in the IMPACC upper respiratory tract data set. **D**. Performance of three-gene classifier with (green) and without (yellow) the addition of age and CT value in the validation cohort. Area under the curve (AUC) listed as value with 95% confidence interval**.**

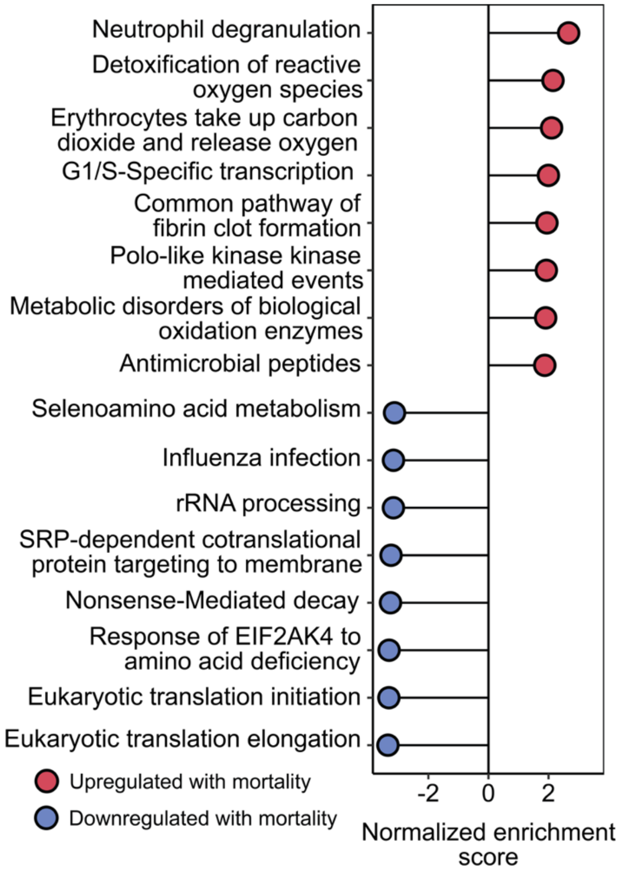


**Figure S2. Pathways associated with mortality in the validation cohort.**
Gene set enrichment analysis (GSEA) demonstrating statistically significant pathways based on Benjamini-Hochberg adjusted p-value in the validation cohort (red = upregulated pathways, blue = downregulated pathways).

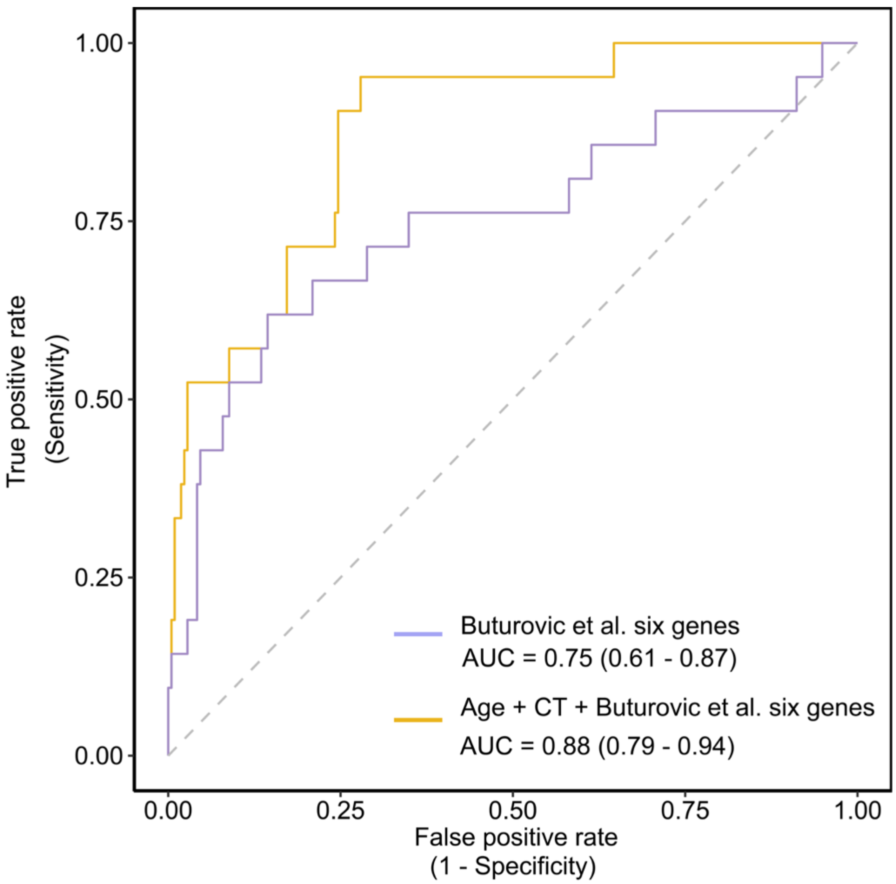


**Figure S3. Performance of a previously published mortality classifier in the IMPACC cohort.** Performance of the Buturovic et al. six gene classifier with (yellow) and without (purple) the added features of age and SARS-CoV-2 cycle threshold in IMPACC. Area under the curve (AUC) listed as value with 95% confidence interval**.**

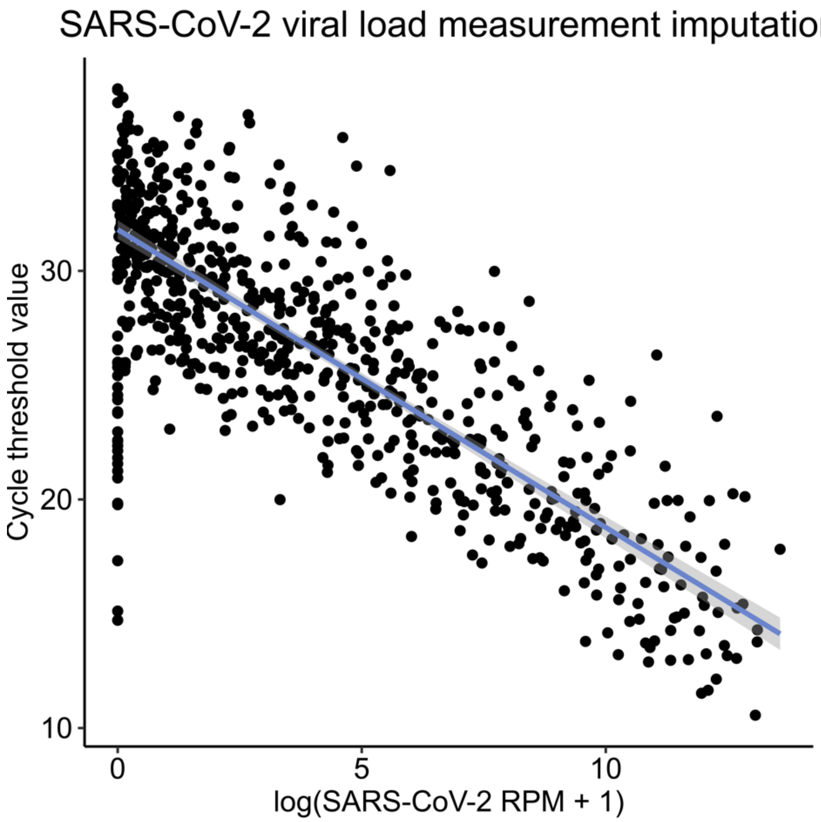


**Figure S4. Robust regression for integrating SARS-CoV-2 reads-per-million (RPM) and cycle threshold values.** Scatter plot showing the relationship between log(SARS-CoV2 RPM + 1) and SARS-CoV-2 cycle threshold (CT), for all subjects with both data available (n = 708). The line of best fit is based on a robust regression model (adjusted R-squared = 0.693, residual standard error = 2.978). Both the intercept and rpM value were significantly associated with CT value (P < 2e-16).

### SUPPLEMENTARY TABLES

| | Survival (n=80) | Mortality (n=42) | P-value |
|---|---|---|---|
| **Age, median (IQR)** | 65.5 (54.0-74.5) | 69.5 (58.0-79.0) | 0.052 |
| **Female, n (%)** | 32 (40.0%) | 14 (33.3%) | 0.557 |
| **Race, n (%)** | | | 0.670 |
| **White** | 34 (42.5%) | 13 (31.0%) | |
| **Black/African American** | 11 (13.8%) | 7 (16.7%) | |
| **Asian** | 22 (27.5%) | 14 (33.3%) | |
| **Other/Declined/Unknown** | 13 (16.3%) | 8 (19.0%) | |
| **Ethnicity** | | | 0.830 |
| **Non-Hispanic** | 68 (85.0%) | 35 (83.3%) | |
| **Hispanic** | 9 (11.3%) | 6 (14.3%) | |
| **Declined/Unknown** | 3 (3.8%) | 1 (2.4%) | |
| **Comorbidities, n (%)** | | | |
| **Diabetes** | 25 (31.3%) | 15 (35.7%) | 0.686 |
| **Hypertension** | 31 (38.8%) | 14 (33.3%) | 0.693 |
| **Chronic pulmonary disease** | 20 (25.0%) | 8 (19.0%) | 0.505 |
| **Cancer** | 13 (16.3%) | 10 (23.8%) | 0.337 |
| **Immunocompromised** | 6 (6.5%) | 7 (16.7%) | 0.133 |
| **Autoimmune disease** | 3 (3.8%) | 0 (0.0%) | 0.0551 |
| **Mechanical ventilation, n (%)** | 65 (81.3%) | 35 (83.3%) | 0.999 |
| **Vasopressor use, n (%)** | 62 (77.5%) | 37 (88.1%) | 0.223 |

**Table S1: Sepsis cohort (EARLI) demographics.** Mann-Whitney test was used for all continuous variables, and Fisher's exact test was used for all categorical values. IQR, interquartile range; EARLI, Early Assessment of Renal and Lung Injury.

| Size | Genes | AUC Mean | AUC Std. Dev. |
|---|---|---|---|
| 2 | ***ATP1B2***, ***DAAM2*** | 0.79 | 0.04 |
| 3 | ***ATP1B2***, ***DAAM2***, ***CD83*** | 0.80 | 0.04 |
| 4 | ***ATP1B2***, ***DAAM2***, *TSPAN7, KCNH8* | 0.81 | 0.04 |
| 5 | ***ATP1B2***, ***DAAM2***, *TSPAN7, KCNH8,* ***CD83*** | 0.82 | 0.04 |
| 6 | ***ATP1B2***, ***DAAM2***, *TSPAN7, KCNH8,* ***CD83***, *FCER1A* | 0.82 | 0.04 |
| 8 | ***ATP1B2***, ***DAAM2***, *TSPAN7, KCNH8,* ***CD83***, *FCER1A, NR4A3, CPA3* | 0.82 | 0.05 |
| 10 | ***ATP1B2***, ***DAAM2***, *TSPAN7, KCNH8,* ***CD83***, *FCER1A, NR4A3, CPA3, TMEM45A, MLNR* | 0.82 | 0.04 |

**Table S2. Peripheral blood candidate mortality classifiers generated by LASSO.** The best performing gene sets for feature lengths n = 2-6, 8, and 10 based on the 3-fold random partitioning evaluation of performance characteristics. Bolded genes are the genes included in the final three gene classifier. LASSO, least absolute shrinkage and selection operator; AUC, area under the receiver operating characteristic curve.

| Length | Genes | AUC mean | AUC Std. Dev |
|---|---|---|---|
| 2 | ***SLC5A5, CD200R1*** | 0.79 | 0.04 |
| 3 | ***SLC5A5, CD200R1**, **FCER1A*** | 0.80 | 0.04 |
| 4 | ***SLC5A5, CD200R1**, **FCER1A**, MARCO* | 0.80 | 0.04 |

**Table S3. Nasal swab candidate mortality classifiers generated by LASSO.** The best performing gene sets for feature lengths n = 2-4 based on the 3-fold random partitioning evaluation of performance characteristics. Bolded genes are the genes included in the final three gene classifier. LASSO, least absolute shrinkage and selection operator; AUC, area under the receiver operating characteristic curve.

| | Survival (n=116) | Mortality (n=21) | P-value |
|---|---|---|---|
| **Age, median (IQR)** | 56.0 (44.3-70.2) | 67.5 (49.9-78.2) | 0.052 |
| **Female, n (%)** | 38 (32.8%) | 8 (38.1%) | 0.625 |
| **Race, n (%)** | | | 0.179 |
| **White** | 38 (32.8%) | 6 (28.6%) | |
| **Black/African American** | 10 (8.6%) | 5 (23.8%) | |
| **Asian** | 18 (15.5%) | 4 (19.0%) | |
| **Other/Declined/Unknown** | 50 (43.1%) | 6 (28.6%) | |
| **Ethnicity, n (%)** | | | 0.822 |
| **Non-Hispanic** | 65 (56.0%) | 13 (61.9%) | |
| **Hispanic** | 48 (41.3%) | 8 (38.1%) | |
| **Declined/Unknown** | 3 (2.6%) | 0 (0.0%) | |
| **Comorbidities, n (%)** | | | |
| **BMI > 30** | 51 (44.0%) | 10 (47.6%) | 0.814 |
| **HIV** | 3 (2.6%) | 1 (4.8%) | 0.490 |
| **Rheumatologic disorder** | 15 (12.9%) | 2 (9.5%) | 0.999 |
| **Solid organ transplant** | 17 (14.7%) | 2 (9.5%) | 0.73 |
| **Mechanical ventilation** | 34 (29.3%) | 16 (76.2%) | <0.001 |
| **Steroid use** | 59 (50.9%) | 17 (81.0%) | 0.016 |
| **Vaccinated for COVID-19** | 45 (38.8%) | 10 (47.6%) | 0.476 |

**Table S4: COVID-19 external validation cohort (COMET) demographics.** Demographics are stratified by survival status. Mann-Whitney test was used for all continuous variables, and Fisher's exact test was used for all categorical values. IQR, interquartile range; BMI, body mass index; COMET, COVID-19 Multi-immunophenotyping projects for Effective Therapies.

### SUPPLEMENTARY DATA FILES

**Supplementary Data File 1**. **A.** Genes differentially expressed with age in IMPACC blood samples (n=785), adjusted for sex and race/ethnicity. **B.** Gene set enrichment analysis (GSEA) of differentially expressed genes. Legend: logFC = log(2) fold change; padj = Benjamini-Hochberg adjusted P value; NES = normalized enrichment score; size = size of Reactome pathway; LeadingEdge = leading edge genes.

**Supplementary Data File 2**. **A.** Genes differentially expressed with age in IMPACC upper respiratory tract samples (n=842), adjusted for sex and race/ethnicity. **B.** Gene set enrichment analysis (GSEA) of differentially expressed genes. Legend: logFC = log(2) fold change; padj = Benjamini-Hochberg adjusted P value; NES = normalized enrichment score; size = size of Reactome pathway; LeadingEdge = leading edge genes.

**Supplementary Data File 3**. **A.** Genes differentially expressed with age in EARLI blood samples (n=122), adjusted for sex and race/ethnicity. **B.** Gene set enrichment analysis (GSEA) of differentially expressed genes. Legend: logFC = log(2) fold change; padj = Benjamini-Hochberg adjusted P value; NES = normalized enrichment score; size = size of Reactome pathway.

**Supplementary Data File 4**. **A.** Genes differentially expressed with age in COMET blood samples (n=137), adjusted for sex and race/ethnicity. **B.** Gene set enrichment analysis (GSEA) of differentially expressed genes. Legend: logFC = log(2) fold change; padj = Benjamini-Hochberg adjusted P value; NES = normalized enrichment score; size = size of Reactome pathway; LeadingEdge = leading edge genes.
